# The Economics of Accuracy for Medical Reasoning with Large Language Models

**DOI:** 10.64898/2025.12.22.25342804

**Authors:** Kiran Bhattacharyya

**Affiliations:** Advanced Product Development, Intuitive Surgical, Inc., Atlanta, GA, USA

## Abstract

Deploying large language models (LLMs) in clinical settings is limited by security, reliability, latency, and accessibility concerns that favor smaller, on-device or on-premise models. However, these smaller models may struggle to meet accuracy requirements. While fine-tuning and retrieval-augmented generation (RAG) can improve domain-specific accuracy, these methods require additional labeled data, technical skill, and infrastructure. In contrast, *test-time scaling* —allocating extra token-budget during inference—offers a training-free alternative to increasing accuracy. However, the trade-offs between these strategies and their interaction with model size remain poorly understood for medical reasoning. To address this gap, we compare three approaches—test-time scaling, fine-tuning, and context grounding—using the Gemma and MedGemma family of LLMs (Gemma-3 1B, Gemma-3 4B, Gemma-3 27B, MedGemma-4B, and MedGemma 27B) and evaluate these systems across common biomedical question-answering (QA) datasets and a set of recently released medical exam questions with the performance of practicing clinicians available for comparison. We test baseline prompts (direct answer, Chain-of-Thought, and self-consistency) while introducing a new prompting method we call “prompt-chaining for continuous reflection” (PCCR) that forces inference time minimum token-generation budgets. We assess accuracy and tokens-generated, allowing us to investigate the accuracy–efficiency trade-offs across prompting, context-grounding, fine-tuning, and model scales. We discover equivalency points where smaller models perform comparably to larger ones with increased reasoning budgets, context-grounding, or fine-tuning. We also find inflection points where context-grounding and test-time scaling used together lead to degrading performance. Using these empirical results, we formulate a general framework with equations to balance cost-benefit trade-offs when engineering LLM-based systems for medical reasoning and QA. We recommend generalizable configurations, designs, and patterns to achieve accuracy and efficiency objectives for example use-cases relevant to healthcare organizations.

**Author summary:** When doctors and hospitals want to use artificial intelligence for medical tasks, they face difficult choices. The most capable AI systems are expensive to run and require sending sensitive patient and hospital data to external servers. Smaller systems that can run locally are more practical but may be less accurate. Our research asked: with what configurations can we make smaller AI systems perform as well as larger ones for medical reasoning? We tested five AI models of different sizes, including both generalist and medically-specialized models, on thousands of medical questions using various prompting strategies, including a new and very simple method we developed that encourages the AI to reason more extensively. We discovered several surprising findings which we describe in detail. Most importantly, we found that smaller, cheaper models can match larger ones when given the right combination of prompting strategy, specialized training, and supporting information. We translated these findings into practical guidelines that can help choose AI configurations that balance accuracy, speed, and cost. Our framework could help make medical AI more accessible to institutions with limited computational resources while achieving the highest possible accuracy.

## Introduction

Large language models (LLMs) have demonstrated remarkable proficiency on medical exams and clinical reasoning tasks achieving scores exceeding 90% on the United States Medical Licensing Examination (USMLE) [1, 2]. However, there are gaps to clinical deployment. Specifically, healthcare organizations must satisfy the systemic constraints of clinical workflows and environments—1) security and privacy requirements that favor on-premise deployment, 2) latency objectives that require timely responses, 3) standards that favor consistency and verifiable outputs, and 4) accessibility requirements that limit computational resources and network bandwidths or availability [3].

However, the dominant paradigm in artificial intelligence (AI) has held that larger models, trained on larger datasets, would yield superior performance—a perspective encapsulated in the neural scaling laws that governed model development [4, 5]. In this view, achieving improvements in clinical AI requires increasing parameter counts to hundreds of billions along with the corresponding massive computational infrastructure, implying acceptance of associated costs, network requirements, and latencies. Current frontier models exemplify this approach, demonstrating impressive performance but requiring enterprise-grade hardware [6] that make clinical systems integrations difficult due to the constraints mentioned previously. On the other hand, three alternative optimization methods have matured that can decouple performance from scale.

First, retrieval-augmented generation (RAG) enables models to access external knowledge at inference time, separating factual knowledge from parametric memory and reducing hallucination rates in medical question-answering (QA) tasks [7–9]. As opposed to a static representation of medical knowledge in model weights that becomes obsolete requiring re-training as clinical guidelines evolve, RAG systems retrieve relevant documents and ground their responses in current or changing evidence and business logic [10–12].

Second, domain-specific parameter-efficient fine-tuning adapts general-purpose models to medical reasoning through exposure to curated clinical datasets [13]. The release of open-weight medical models—including Med-Llama, Meditron, and MedGemma—has demonstrated that medically-adapted LLMs with smaller sizes can show promising performance on medical QA and reasoning tasks [14, 15]. However, this approach has limitations since fine-tuned models are “locked” into the knowledge state of their training data, and the process of curating expert-reviewed medical training sets and fine-tuning models requires significant expertise and resources [16].

Third, test-time scaling has emerged as a training-free alternative for enhancing model reasoning. Popularized by models such as OpenAI’s o1 and frameworks such as the “m1” and “s1” [17, 18], test-time scaling allocates additional computational budget during inference rather than training. By inducing the model to generate extended chains of reasoning, smaller models can achieve accuracy levels comparable with their larger counterparts. Research has demonstrated that a 32-billion parameter model, when afforded a reasoning budget of approximately 4,000 tokens, can match the performance of much larger models on complex math reasoning tasks [18]. This suggests a fungibility between model parameters and inference computation with implications for deployment economics [19].

Despite the promise of these approaches, their interactions and trade-offs across model sizes remain poorly characterized, particularly in medical contexts. Several critical questions lack empirical grounding: How does the benefit of chain-of-thought prompting vary with model scale? At what point does extended reasoning become counterproductive—causing models to “overthink” and second-guess correct initial responses? How do the accuracy gains from fine-tuning interact with sophisticated prompting strategies? When relevant context is available, does additional reasoning help or hinder performance? And perhaps most practically: given specific accuracy requirements, latency constraints, and computational budgets, which combination of model size, fine-tuning, retrieval augmentation, and reasoning budget offers the optimal trade-off?

These questions have practical implications for healthcare organizations who face concrete decisions with significant resource implications for capital and operational expenses. The choice between investing in RAG infrastructure versus fine-tuning workflows versus simply purchasing more inference compute depends critically on understanding how these approaches interact across different clinical use cases—from real-time emergency department triage requiring sub-second responses to asynchronous rare disease consultation where sixty-second latencies are acceptable.

Here, we address this knowledge gap through a systematic empirical investigation of the Gemma and MedGemma model families across medical reasoning tasks [15, 20]. We selected this family of models for several reasons. First, Gemma models span a range of sizes (1B, 4B, and 27B parameters) accessible to computational environments ranging from mobile devices to single consumer GPUs. Second, the existence of base models (Gemma-3) and medically fine-tuned variants (MedGemma) derived from identical architectures and checkpoints enables direct measurement of fine-tuning effects without confounding architectural differences. Third, the medically fine-tuned model variants represent the current state of open-weight LLMs, with MedGemma 27B reporting 87.7% on MedQA which is comparable with frontier closed models while operating at a much lower inference cost [15].

We evaluate these models across three medical QA datasets with multiple-choice answers spanning different reasoning demands and knowledge requirements. We test the models internalized medical knowledge with MIRAGE which provides a large-scale evaluation across different consolidated biomedical datasets [21]. On the other hand, PubMedQA includes “oracle” context, enabling assessment of how models utilize external evidence and whether additional reasoning helps or hinders when relevant information is provided [22]. Finally, we incorporate a set of recently released medical licensing examination questions that includes human physician performance benchmarks, allowing direct comparison to clinician accuracy [23].

We perform a systematic comparison of prompting strategies along the test-time scaling continuum. We evaluate baseline approaches: direct answering (P0), single-pass chain-of-thought (P1) [24, 25], and self-consistency with majority voting across five samples (P2) [26–28]. We additionally introduce a new prompting method we term “prompt-chaining for continuous reflection” (PCCR), which enforces minimum token-generation budgets through iterative prompting similar to s1 test-time scaling [18] but PCCR is usable over API-level access to LLMs.

Our findings provide several key insights and the empirical grounding needed to make practical decisions about LLM deployment in healthcare settings. First, we demonstrate that chain-of-thought prompting has the expected effect on larger models (27B) but mixed performance for smaller models (1B-4B)–in some cases, showing a 6.9% decrease in accuracy when prompted to reason step-by-step compared to direct answering. Second, we identify a nuanced interaction between test-time scaling and context-grounding where certain prompting methods can lead to “overthinking” that varies by task and context availability. Third, we find that fine-tuning and sophisticated prompting exhibit strong positive interaction effects—the accuracy advantage of MedGemma over base Gemma grows from 4.6% with direct prompting to 15.7% percentage points with self-consistency voting—suggesting that the value of medical fine-tuning is substantially underestimated when evaluated only with simple prompting strategies. Finally, we find that no model-prompt configuration receives a passing score on the medical exam dataset but the performance overlaps with the lowest quartile of practicing physicians.

These empirical findings enable us to formulate a general framework for reasoning about LLM deployment trade-offs in medical settings. We develop equations relating accuracy to model scale, fine-tuning status, prompting strategy, reasoning budget, and context availability, with parameters inferred from our experimental data. We derive cost and latency models that account for the computational overhead of extended reasoning and retrieval augmentation. Using this framework, we identify equivalence points—configurations where smaller models match larger model performance through compensating strategies—and inflection points where combining approaches leads to degraded rather than improved outcomes. Finally, we translate these findings into practical recommendations, presenting decision frameworks and architectural patterns for common healthcare deployment scenarios.

## 1 Methods

We conducted a systematic evaluation of five language models from the Gemma and MedGemma families across three medical question-answering datasets using four prompting strategies. Our experimental design was structured to investigate the effects of model size, medical fine-tuning, test-time scaling, and context grounding on accuracy and efficiency. All experiments were conducted using consistent evaluation infrastructure to ensure reproducibility. Generations were evaluated using a ‘LLM-as-judge’ which was validated with manual review with a set of 200 QA pairs. We report accuracy and estimate 95% confidence intervals (CIs) with the binomial approximation method. We inflate the CIs to account for imperfect adjudication from the LLM-as-judge, as explained later.

### 1.1 Models

We evaluated five models spanning three parameter scales, with matched pairs of base and medically fine-tuned variants at the 4B and 27B scales. All evaluations used in this study were text-only. However, it is important to note that some of the models below support text and image inputs.

- **Gemma-3 1B.** The smallest model in our evaluation, Gemma-3 1B is a text-only decoder transformer with approximately 1 billion parameters. This model represents the lower bound of contemporary LLM capability, suitable for deployment on mobile devices. We include this model to characterize performance floors and assess whether test-time scaling can compensate for limited model capacity.
- **Gemma-3 4B.** A multimodal vision-language model with approximately 4 billion parameters, Gemma-3 4B employs a decoder-only transformer architecture with a

SigLIP vision encoder. For our text-only evaluation, we utilized the model without image inputs.

- **MedGemma 4B.** A medical adaptation of Gemma-3 4B. MedGemma 4B was fine-tuned on medical text and image data including radiology images, histopathology slides, ophthalmology images, and dermatology images. The architectural specifications are identical to Gemma-3 4B with an updated vision-encoder. However, our text-only inputs are not processed by this updated vision-encoder.
- **Gemma-3 27B.** This is the largest base model in our evaluation. Gemma-3 27B is a multimodal vision-language model containing approximately 27 billion parameters and was trained on 10+ trillion tokens including web documents, code, and mathematical text in 100+ languages.
- **MedGemma 27B.** The medically fine-tuned variant of Gemma-3 27B, optimized for text-only medical reasoning tasks.

Extensive technical details can be found for all models above in the Gemma-3 and MedGemma technical reports [15, 20]. All models were self-hosted on A100 GPUs using the HuggingFace transformers library [29] using Python on a Linux system running Ubuntu 24.04. We used BF16 precision for all evaluations. Temperature was set to 0.2 for more deterministic prompting conditions (P0, P1, PCCR) and 0.7 for self-consistency sampling (P2).

### 1.2 Datasets

We evaluated models on three medical question-answering datasets selected to span different reasoning demands, knowledge sources, and clinical relevance.

- **MIRAGE.** This dataset comprises 7,663 multiple-choice questions aggregated from diverse biomedical question-answering sources [21]. Questions span clinical medicine, basic science, pharmacology, and pathophysiology. This dataset includes 1) 1,089 QA from MMLU-Med, 2) 1,273 QA from MedQA-US, 3) 4,183 QA from MedMCQA, 4) 500 QA from PubMedQA, and 5) 618 questions from BioASQ-Y/N. Critically, we evaluated models on MIRAGE *without providing any supporting context*, requiring models to rely entirely on parametric knowledge.
- **PubMedQA.** A benchmark dataset comprising 1,000 QA pairs derived from PubMed abstracts, where each question can be answered using the context in the corresponding abstract [22]. We evaluated models under two conditions: (1) without context, requiring the model to answer based solely on parametric knowledge, and (2) with oracle context, where the relevant PubMed abstract was provided in the prompt. This paired design enables direct measurement of context grounding effects and assessment of how test-time scaling interacts with retrieved evidence. The oracle context represents an upper bound, which removes confounding effects of variable retrieval quality.
- **Medical Licensing Examination Questions.** This publicly-available dataset comprises 655 multiple-choice questions from five different specialties: internal medicine, general surgery, pediatrics, psychiatry, and obstetrics/gynecology [23]. Importantly, this dataset includes benchmark performance data from 849 practicing clinicians and a threshold for passing scores, enabling direct comparison between model and human physician accuracy. Questions were presented without any supporting context.

All datasets employed multiple-choice formats ranging from 2–4 answer options depending on the dataset and the question. We extracted model responses using a combination of constrained generation, regex parsing, and LLM-as-judge. Questions where the model failed to produce a valid answer selection were scored as incorrect.

### 1.3 Prompting strategies

We evaluated four prompting strategies representing increasing levels of test-time computation. We describe each prompting pattern here and provide the full prompts in Appendix A.

- **P0: Direct Answering.** The baseline condition in which models were prompted to select an answer without any reasoning. This configuration minimizes token generation and inference latency, representing the lower bound of test-time computation.
- **P1: Chain-of-Thought.** Models were prompted to reason step-by-step before providing a final answer, following established chain-of-thought methodology [24, 25]. This approach has been shown to improve performance on complex reasoning tasks for sufficiently large models, though its effects on smaller models are less well characterized.
- **P2: Self-Consistency with Majority Voting.** Following [26, 27], we sampled five independent chain-of-thought responses (k=5) at temperature 0.7 and selected the final answer by majority vote. When ties occurred, we used random selection. This approach leverages the observation that correct reasoning paths are more likely to converge on consistent answers, while errors tend to be more variable.
- **Prompt-Chaining for Continuous Reflection (PCCR).** We introduce a novel prompting strategy designed to enforce minimum reasoning budgets through iterative prompting. The procedure operates as follows:

1. The model receives an initial prompt requesting chain-of-thought reasoning (identical to P1) and generates a response.
2. If the chain-of-thought portion meets or exceeds the specified token budget, the response is accepted and the final answer is extracted.
3. If the chain-of-thought falls below the token budget, the system constructs a continuation prompt that includes the original question, all previously generated reasoning, and an instruction to continue reflecting on the problem.
4. Steps 2–3 repeat, accumulating reasoning across iterations, until the combined chain-of-thought reaches or exceeds the target budget.
5. The final answer is extracted from the terminal response.

We evaluated PCCR at five token budget levels: 128, 256, 512, 768, and 1024 tokens.

These budgets refer to the minimum total reasoning tokens accumulated across all continuation iterations, not including the original question or final answer tokens. Token counts were approximated by word count. This approach differs from prior test-time scaling methods in several respects. Unlike budget forcing through “Wait” token injection [18], PCCR operates through natural language continuation instructions, making it compatible with any instruction-following model without specialized training or direct intervention in the token generation process—making it accessible over APIs. Finally, unlike simple extended generation, PCCR ensures that the model explicitly revisits and reflects on its prior reasoning, potentially enabling error correction and consideration of alternative hypotheses.

### 1.4 Evaluation

#### 1.4.1 Automated Accuracy Assessment

We used constrained generation and regex to extract the final answer from the LLM response. If this extraction resulted in an empty string, the answer was scored as incorrect. In many cases, the extracted answer would not have the multiple choice label (A, B, C, etc.) but a full text answer that mapped to one of the choices along with a longer explanation. For this reason, we measured accuracy using a *LLM-as-judge* (Mistral Small 3.1 24B). The judge was prompted to evaluate if a generated answer was factually consistent, complete, and relevant compared to the ground-truth answer which was a multiple choice answer (full prompt in the Appendix B). To ensure reliability, the prompt included three few-shot examples of correct and incorrect evaluations.

### Human Validation of the LLM-as-judge

The validity of using an LLM-as-judge is critical for the credibility of our results. We conducted a blinded human evaluation on 200 randomly sampled model outputs (100 judged as correct, 100 incorrect). A human evaluator, using the same criteria, showed a high degree of agreement with the LLM-as-judge, achieving a Cohen’s *κ* score of 0.99 (Table 1). This strong correlation provides confidence in our automated evaluation pipeline.

**Table 1.** Confusion matrix validating the LLM-as-judge against human evaluation for 200 QA pairs.

| Human Label | LLM: Correct | LLM: Incorrect |
| --- | --- | --- |
| Correct | 99 | 1 |
| Incorrect | 1 | 99 |

Our primary metric was multiple-choice accuracy, calculated as the proportion of questions for which the model selected the correct answer. We report accuracy as percentages with 95% confidence intervals (CI_95%_) calculated using the normal approximation to the binomial distribution described in Equation 1, where *p*^ is the observed accuracy and *n* is the number questions in the QA dataset.

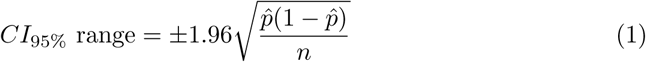

To account for residual labeling uncertainty in the LLM-as-judge (Cohen’s *κ* = 0.99 with human raters), we generated 10,000 bootstrapped samples by pseudo-randomly flipping correct and incorrect answers with the corresponding error-rate of the LLM-as-judge (1%) for each model-prompt configuration for each dataset. We then used these 10,000 uncertainty-aware estimates of *p*^ to compute 10,000 CI_95%_ intervals from which we chose the largest interval. This expansion of CIs often made them comparable across the ranges of reported *p*^ values (∼0.2-0.8) for a given *n*. Therefore, *we report one CI range (the largest) per QA dataset* that applies to all *p*^ values.

### 1.5 Statistical analyses

We use McNemar’s test for paired binary outcomes when comparing the performance of 2 model-prompting configurations (e.g., performance of MedGemma 27B with P0 vs Gemma 27B with P1). We use the term “percentage points” or “Δ%” when discussing differences between or ranges in accuracy of model-prompting configurations reported in percent (%). To account for the imperfect adjudication of the LLM-as-judge when performing statistical tests, we only compare model-prompt configurations with accuracy values that are at least one expanded, full CI_95%_ range away from each other. For this reason, *p*-values were generally quite small (10^−3^ − 10^−300^) since dataset sizes were fairly large 655 − 7663. We report *p*-values at orders of magnitude (ie, *p <* 10^−3^ or *p <* 10^−4^) and stop reporting differences in *p*-value after 10^−5^ as the practical meaning of these minute differences are unclear.

### 1.6 Reproducibility

All code and data is available through this GitHub repo. https://github.com/MiningMyBusiness/accuracy-econ-medical-llms

## 2 Results

We evaluated five models from the Gemma and MedGemma families across three medical question-answering datasets using four prompting strategies. Our results reveal consistent patterns regarding the effects of model scale, medical fine-tuning, test-time scaling, and context grounding on accuracy. We first present the empirical findings organized by dataset, inspect specific characteristics of responses test-time scaling approaches, then synthesize these observations into a general framework for reasoning about deployment trade-offs.

### 2.1 Effects of Prompting and Model Scale on MIRAGE

Table 2 presents accuracy results across all model and prompting combinations on the MIRAGE dataset (N = 7,663 questions, no context provided). Below, we highlight key patterns that emerge from these results.

**Table 2.**
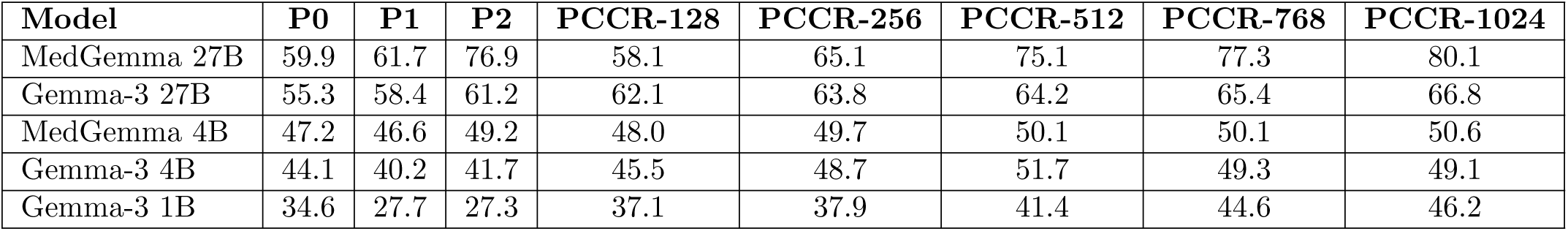
Accuracy (%) on MIRAGE dataset by model and prompting strategy. N = 7,663 questions, no supporting context provided. CI_95%_ is ±1.2 percentage points for all values.

#### 2.1.1 Chain-of-thought (P1) can harm smaller models

Contrary to the assumption that explicit reasoning universally improves performance, we observed that chain-of-thought prompting (P1) decreased accuracy relative to direct answering (P0) for smaller models. Gemma-3 1B showed a 6.9 Δ% decrease (34.6% to 27.7%, McNemar’s test, *p <* 10^−4^), and Gemma-3 4B showed a 3.9 Δ% decrease (44.1% to 40.2%, McNemar’s test, *p <* 10^−3^). MedGemma 4B performance difference was within the CI_95%_ (±1.2 percentage points) for P0 and P1. In contrast, the Gemma-3 27B model showed improvements with P1 (+3.1 Δ%, *p <* 10^−3^). MedGemma 27B improved more modestly by 1.8 Δ% with partially overlapping CI_95%_. These results suggest that chain-of-thought reasoning requires sufficient model size to execute coherently; smaller models may may not benefit from it.

#### 2.1.2 Self-consistency (P2) shows model dependent gains

Majority voting across five chain-of-thought samples (P2) produced notable accuracy improvements for MedGemma 27B (+15.2 Δ% over P1, from 61.7% to 76.9%, McNemar’s test, *p <* 10^−5^). However, gains were markedly smaller for Gemma-3 27B (+2.8 Δ%, *p <* 10^−2^) and MedGemma 4B (+2.6 Δ%, *p <* 10^−2^). Differences were minimal or absent for the Gemma-3 4B and 1B models. This pattern indicates that self-consistency is most effective when the underlying model produces high-quality but variable reasoning chains—a condition that may be better satisfied by larger or fine-tuned models.

#### 2.1.3 PCCR enables accuracy improvements with extended reasoning

Our prompt-chaining for continuous reflection method (PCCR) demonstrated increasing accuracy with reasoning budget for MedGemma 27B and Gemma-3 27B on this dataset (Figure 1A). Accuracy improved from 58.1% at 128 tokens to 80.1% at 1024 tokens for MedGemma 27B—a gain of 22.0 Δ% (McNemar’s test, *p <* 10^−5^). Notably, PCCR-1024 (80.1%) exceeded even P2 (76.9%) by 3.2 Δ% (*p <* 10^−2^), suggesting that enforced extended reasoning can outperform sampling-based approaches. Gemma-3 27B showed more modest but consistent gains with PCCR, improving from 62.1% to 66.8% (McNemar’s test, *p <* 10^−3^) across the same budget range, also showing a large difference between PCCR-1024 and P2 (+5.6 Δ%, *p <* 10^−4^).

**Fig 1.**
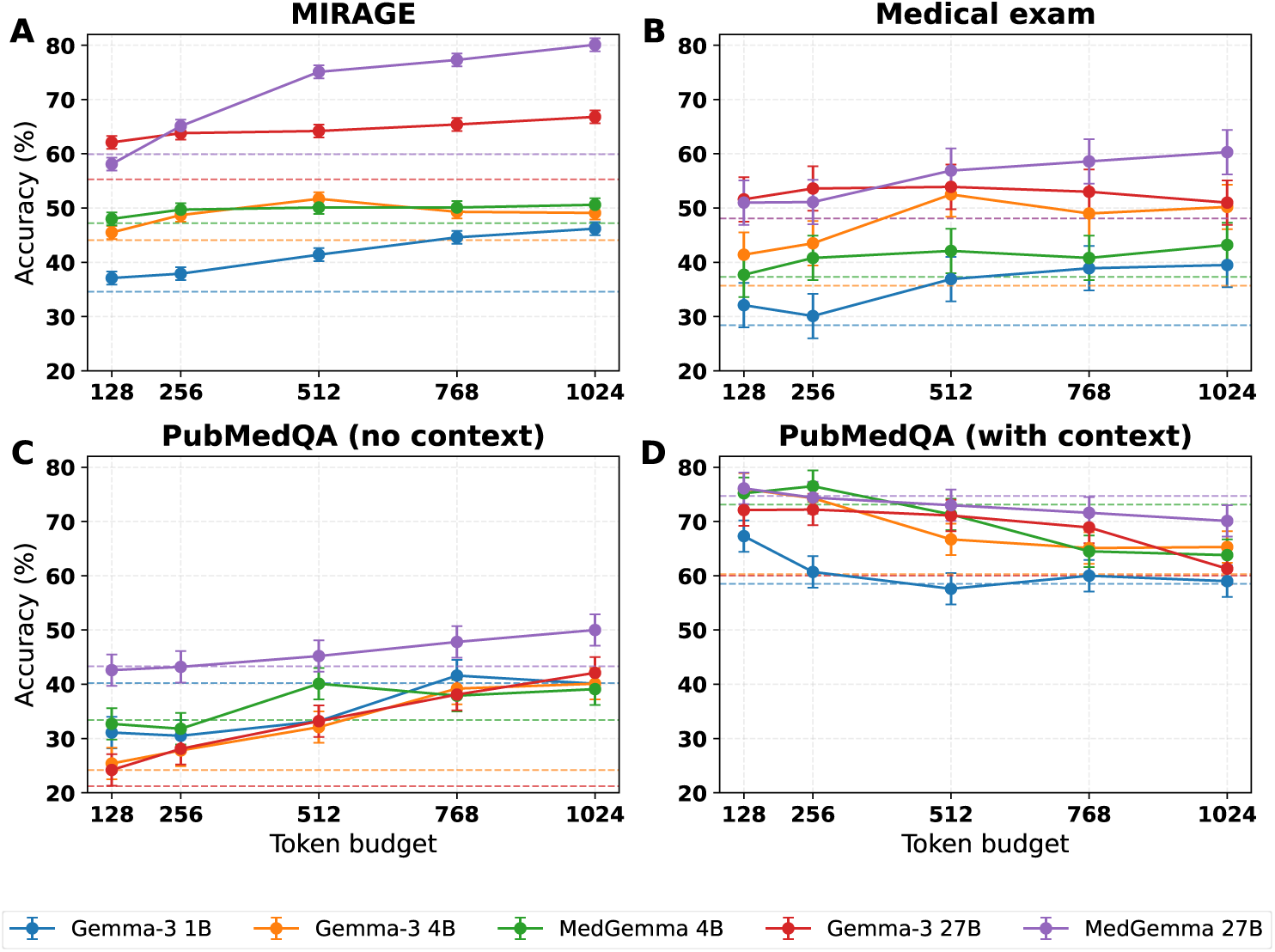
Accuracy as a function of PCCR token budget across models and datasets. Each panel shows accuracy (%) versus minimum reasoning token budget for the five evaluated models. Error bars indicate 95% confidence intervals (refer to Section 1.4.1 for details). Dashed lines in each sub-figure are reference lines of model performances for the direct-answer prompting method (P0). Top row: datasets without supporting context (MIRAGE, N=7,663; Medical exam, N=655). Bottom row: PubMedQA (N=1,000) evaluated without context (C) and with oracle context (D). Without context (A, B, & C), accuracy generally increases with token budget, with MedGemma 27B showing the largest gains. With oracle context (D), accuracy peaks at low token budgets (128–256 tokens) and declines with extended reasoning.

#### 2.1.4 Smaller models benefit from PCCR but differently

Gemma-3 1B showed substantial improvement with PCCR, increasing from 37.1% at 128 tokens to 46.2% at 1024 tokens (+9.1 Δ%, McNemar’s test, *p <* 10^−5^). This improvement exceeded the model’s P0 baseline (34.6%) by 11.6 Δ%, partially compensating for the model’s limited capacity. However, the 4B models showed ceiling effects, with accuracy plateauing around 50-51% regardless of extended reasoning budget. Using PCCR, the 1B model reached comparable performance to P0 prompting for 4B models, but neither 1B nor 4B models could reach P0 performance of 27B models (Figure 1A).

#### 2.1.5 Fine-tuning amplifies the benefits of prompting

Table 3 presents the accuracy difference of MedGemma over the corresponding base Gemma model at each prompting strategy, revealing an interaction effect. At the 27B size, the fine-tuning advantage grew from +4.6 Δ% with direct answering (P0) to +15.7 Δ% with self-consistency (P2), representing a 3.4-fold amplification of the fine-tuning benefit. Similarly, PCCR-1024 showed a +13.3 Δ% advantage compared to +4.6 Δ% for P0. The 4B scale showed a similar but weaker pattern. These findings indicate that combining medical fine-tuning with sophisticated prompting strategies may amplify accuracy improvements over the base model.

**Table 3.** Fine-tuning benefit on MIRAGE (MedGemma accuracy – Gemma-3 accuracy) by model size and prompting strategy on MIRAGE dataset. Values in percentage points.

| Scale | P0 | P1 | P2 | PCCR-128 | PCCR-256 | PCCR-512 | PCCR-768 | PCCR-1024 |
| --- | --- | --- | --- | --- | --- | --- | --- | --- |
| 27B | +4.6 | +3.3 | +15.7 | -4.0 | +1.3 | +10.9 | +11.9 | +13.3 |
| 4B | +3.1 | +6.4 | +7.5 | +2.5 | +1.0 | -1.6 | +0.8 | +1.5 |

### 2.2 Effect of Context Grounding on PubMedQA

Table 4 presents accuracy on PubMedQA under two conditions: without context and with oracle context—providing the relevant PubMed abstract. This design enables direct assessment of how context grounding interacts with model scale and prompting strategy.

**Table 4.** Accuracy (%) on PubMedQA dataset. Larger values indicate accuracy with oracle context; values in parentheses indicate accuracy without context. N = 1,000 questions. 95% confidence interval is ±2.9 percentage points.

| Model | P0 | P1 | P2 | PCCR-128 | PCCR-256 | PCCR-512 | PCCR-768 | PCCR-1024 |
| --- | --- | --- | --- | --- | --- | --- | --- | --- |
| MedGemma 27B | (43.3) 74.7 | (35.3) 77.4 | (42.1) 75.2 | (42.6) 76.1 | (43.2) 74.4 | (45.2) 73.0 | (47.8) 71.6 | (50.0) 70.1 |
| Gemma-3 27B | (21.2) 60.0 | (18.1) 68.4 | (22.3) 70.1 | (24.2) 72.1 | (28.1) 72.2 | (33.2) 71.1 | (38.1) 68.9 | (42.1) 61.3 |
| MedGemma 4B | (33.4) 73.1 | (24.2) 56.2 | (32.5) 74.3 | (32.7) 75.2 | (31.8) 76.5 | (40.1) 71.3 | (37.9) 64.5 | (39.1) 63.8 |
| Gemma-3 4B | (24.2) 60.3 | (20.1) 64.7 | (23.9) 70.1 | (25.4) 76.0 | (27.8) 74.3 | (32.1) 66.7 | (39.2) 65.1 | (40.1) 65.3 |
| Gemma-3 1B | (40.2) 58.5 | (18.0) 58.6 | (33.7) 60.1 | (31.1) 67.3 | (30.5) 60.7 | (33.2) 57.6 | (41.6) 60.0 | (40.1) 59.0 |

Across all models and prompting strategies, providing oracle context improved accuracy substantially. For MedGemma 27B with direct answering (P0), accuracy increased from 43.3% to 74.7%—a gain of 31.4 percentage points (McNemar’s test, *p <* 10^−5^). Even Gemma-3 1B improved from 40.2% to 58.5% (+18.3 percentage points, *p <* 10^−5^). These gains demonstrate the value of retrieval-augmented generation for medical question-answering.

#### 2.2.1 Chain-of-thought without context degrades performance for most models

Consistent with findings on MIRAGE, P1 prompting without context for PubMedQA reduced accuracy compared to P0 for most models (Table 4). MedGemma 27B dropped from 43.3% to 35.3% (−8.0 Δ%, *p <* 10^−3^), Gemma-3 27B from 21.2% to 18.1%, and most dramatically, Gemma-3 1B from 40.2% to 18.0% (−22.2 Δ%, *p <* 10^−5^). MedGemma 4B showed a similar pattern, dropping from 33.4% to 24.2% (−9.2 Δ%, *p <* 10^−3^). These results reinforce the finding that chain-of-thought prompting can be counterproductive when models lack sufficient knowledge or capacity, causing them to reason their way to incorrect answers.

#### 2.2.2 Self-consistency (P2) provides robust gains with context across all model scales

A notable finding emerged from the P2 results for PubMedQA (Table 4): majority voting produced consistent accuracy improvements with oracle context across all models, including smaller ones that showed minimal P2 benefit on MIRAGE. Table 5 presents the P2 gains relative to P0 and P1 for each model.

**Table 5.** Self-consistency (P2) performance relative to other prompting strategies on PubMedQA with oracle context.

| Model | P0 | P1 | P2 | P2 – P0 ($\Delta\%$) | P2 – P1 ($\Delta\%$) |
| --- | --- | --- | --- | --- | --- |
| MedGemma 27B | 74.7% | 77.4% | 75.2% | +0.5 | -2.2 |
| Gemma-3 27B | 60.0% | 68.4% | 70.1% | +10.1 | +1.7 |
| MedGemma 4B | 73.1% | 56.2% | 74.3% | +1.2 | +18.1 |
| Gemma-3 4B | 60.3% | 64.7% | 70.1% | +9.8 | +5.4 |
| Gemma-3 1B | 58.5% | 58.6% | 63.1% | +4.6 | +4.5 |

For the base Gemma models, P2 with context achieved the highest or near-highest accuracy among all prompting strategies: Gemma-3 27B reached 70.1% (comparable to its PCCR-128 performance), Gemma-3 4B reached 70.1% (nearly overlapping CIs with PCCR-128 at 76.0%), and Gemma-3 1B reached 63.1% (overlapping CIs with PCCR-128 at 67.3%). This pattern suggests that self-consistency voting is particularly effective when models have access to relevant context but may produce variable reasoning chains—voting filters out erroneous reasoning while preserving correct extractions from the provided evidence.

MedGemma 4B exhibited anomalous behavior with P1 prompting: accuracy with context dropped from 73.1% (P0) to 56.2% (P1)—a 16.9 percentage point degradation (*p <* 10^−5^) that was the largest P0-to-P1 drop observed in any condition. Remarkably, P2 recovered this performance entirely, achieving 74.3%—slightly exceeding P0 but within confidence intervals. This recovery suggests that MedGemma 4B’s P1 failures stemmed from high-variance reasoning rather than systematic error; majority voting may have filtered out the erroneous reasoning chains while preserving correct ones.

For high-performing configurations, P2 showed some ceiling effects. For instance, MedGemma 27B, which already achieved strong performance with P1 (77.4%), P2 provided no additional benefit and showed slight degradation to 75.2% (−2.2 Δ%, within confidence intervals). This suggests that when a model’s reasoning is already reliable, the overhead of multiple samples provides no benefit and may introduce noise.

#### 2.2.3 Extended reasoning degrades performance when context is available

A repeating trend emerged when comparing PCCR performance with and without context. Without context, extended reasoning generally improved accuracy (Figure 1C). However, with oracle context, extended reasoning beyond short budgets degraded performance (Figure 1D). For MedGemma 27B, accuracy peaked at P1 (77.4%) and declined monotonically with increasing PCCR budget, reaching 70.1% at PCCR-1024—a loss of 7.3 Δ% from the optimum (McNemar’s test, *p <* 10^−2^). This pattern was consistent across models.

These results in Table 6 reveal an “overthinking penalty” that varies by model when using context-grounded generation with extended reasoning: when high-quality context is provided, the model’s optimal strategy is to extract and apply the relevant information with the appropriate additional reasoning. Extended reflection leads the model to second-guess correct context-derived answers, introducing errors. We investigate this “overthinking” penalty from PCCR with context further in Section 2.5.

**Table 6.**
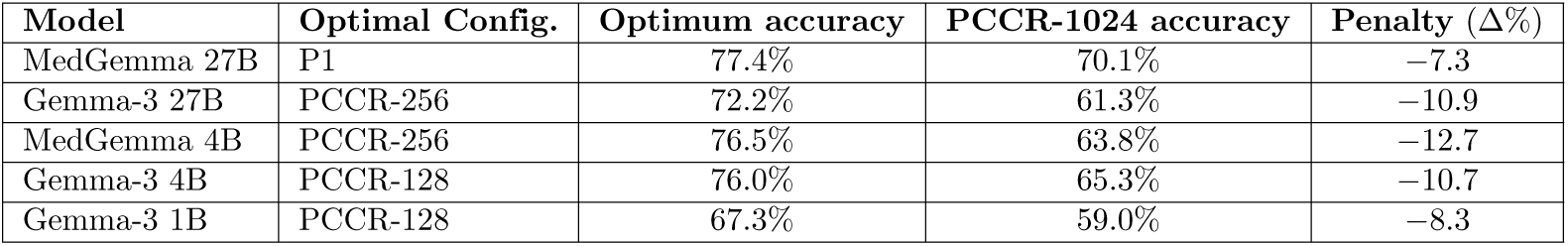
Optimal reasoning budget and “overthinking penalty” with oracle context on PubMedQA. Overthinking penalty calculated as accuracy at PCCR-1024 minus accuracy at optimal configuration.

#### 2.2.4 Divergent effects of P2 versus PCCR reveal distinct mechanisms

The contrasting behavior of P2 and PCCR with oracle context reveals important mechanistic differences between these test-time scaling approaches. P2 (self-consistency) samples multiple independent reasoning chains and aggregates via voting, while PCCR accumulates reasoning within a single extended chain. When high-quality context is available, PCCR’s extended sequential reasoning leads models to second-guess correct context-derived answers—an “overthinking” effect that compounds across continuation rounds. In contrast, P2’s independent sampling produces some chains that correctly extract information from context and others that overthink; majority voting preferentially selects the correct answers, providing a form of robustness against overthinking.

This distinction is illustrated by comparing MedGemma 27B across strategies with context: P1 achieves 77.4%, P2 achieves 75.2% (slight degradation within confidence intervals), while PCCR-1024 achieves only 70.1%. The 5× token cost of P2 relative to P1 yields no benefit but also no harm, whereas the similar token cost of PCCR-1024 yields substantial harm. We examine the effect of P2 on LLM response variability further in Section 2.4.

#### 2.2.5 Without context, P2 shows limited benefit and PCCR shows gains

The pattern reverses when context is unavailable. Without oracle context, P2 provided minimal benefit for most models: MedGemma 27B showed essentially no change (43.3% P0→42.1% P2), and gains for other models were modest. In contrast, PCCR showed consistent accuracy improvements without context, with MedGemma 27B improving from 43.3% (P0) to 50.0% (PCCR-1024, *p <* 10^−3^). This pattern suggests that extended reasoning is valuable when models must rely on parametric knowledge but counterproductive when relevant information is externally provided. This is clearly visible when comparing Figure 1C & D.

#### 2.2.6 Small models with context can match larger models

Comparing across model scales with oracle context reveals notable equivalence points in Table 4. Gemma-3 4B with PCCR-128 achieved 76.0% accuracy, exceeding MedGemma 27B with P0 (74.7%) by 1.3 percentage points (difference within CI_95%_). Similarly, MedGemma 4B with PCCR-256 achieved 76.5%, matching MedGemma 27B P1 (77.4%) within confidence intervals. These equivalences demonstrate that effective retrieval combined with appropriate prompting can substitute for substantial model scale.

### 2.3 Performance on Medical Licensing Examination

Table 7 presents accuracy on the medical licensing examination dataset, which enables comparison with practicing clinician performance. All models showed reduced accuracy on the medical examination compared to MIRAGE (Table 2) which was also tested without context, suggesting this dataset presents greater difficulty. With PCCR, performance on the medical exam dataset approaches P0 performances in MIRAGE (Figure 1A & B).

**Table 7.** Accuracy (%) on medical licensing examination questions. N = 655 questions, no supporting context provided. 95% confidence interval is ±4.1 percentage points.

| Model | P0 | P1 | P2 | PCCR-128 | PCCR-256 | PCCR-512 | PCCR-768 | PCCR-1024 |
| --- | --- | --- | --- | --- | --- | --- | --- | --- |
| MedGemma 27B | 48.1 | 54.5 | 57.3 | 51.0 | 51.1 | 56.9 | 58.6 | 60.3 |
| Gemma-3 27B | 48.1 | 53.6 | 52.1 | 51.6 | 53.6 | 53.9 | 53.0 | 51.0 |
| MedGemma 4B | 37.3 | 36.9 | 37.0 | 37.7 | 40.8 | 42.1 | 40.8 | 43.2 |
| Gemma-3 4B | 35.7 | 34.0 | 37.6 | 41.4 | 43.5 | 52.5 | 49.0 | 50.2 |
| Gemma-3 1B | 28.4 | 21.5 | 31.2 | 32.1 | 30.1 | 36.9 | 38.9 | 39.5 |

The detrimental effect of chain-of-thought on small models replicated: Gemma-3 1B dropped from 28.4% (P0) to 21.5% (P1), a 6.9 Δ% decrease (within CIs). MedGemma 27B again showed the strongest performance with extended reasoning, reaching 60.3% at PCCR-1024. Unlike on MIRAGE, Gemma-3 4B showed substantial improvement with PCCR on this dataset, reaching 52.5% at PCCR-512—higher than MedGemma 4B’s best performance (43.2% at PCCR-1024, *p <* 10^−2^). This reversal of the expected fine-tuning advantage warrants further investigation. For Gemma-3 4B, MedGemma 4B, and Gemma-3 27B, we see some plateauing effects with extended PCCR reasoning where performance stays within CI_95%_ from PCCR-512 onward.

All models perform below the passing score of 65% and below the median physician scores which ranges from 70%-80% depending on the specialty [23]. However, the scores for models did overlap with the bottom quartile of physicians, who had performance ranges from 40%-65% across specialties (range inferred from Fig. 2 in [23]).

**Fig 2.**
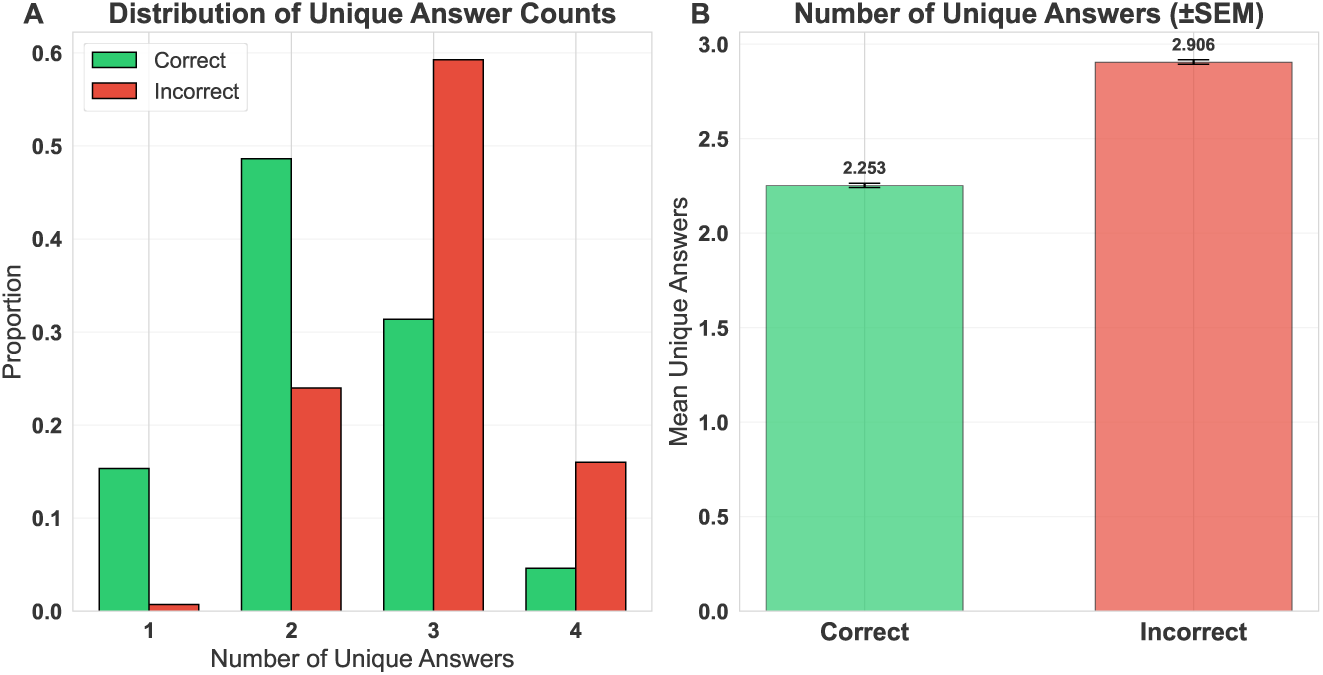
Answer variability in self-consistency (P2) sampling predicts response correctness. Analysis of 7,200 four-option multiple-choice questions across all datasets and models. For each question, five independent chain-of-thought responses were sampled and the number of unique answer options selected was counted. (A) Distribution of unique answer counts, stratified by whether the majority-voted final answer was correct (green) or incorrect (red). Correct responses show higher concentration at lower variability (1–2 unique answers), while incorrect responses are skewed toward higher variability (3–4 unique answers). (B) Mean number of unique answers by correctness. Correct responses averaged 2.25 unique answers compared to 2.91 for incorrect responses (Cohen’s *d* = 0.90, *p <* 10^−5^, Mann-Whitney U test). Error bars indicate standard error of the mean.

### 2.4 Self-Consistency (P2) Agreement as a Calibration Signal

To investigate whether self-consistency voting provides information beyond the final answer, we analyzed the variability of responses within each P2 sample set. For each of the 7,200 four-option multiple-choice questions evaluated across all datasets and models, we counted the number of unique answer options selected among the five sampled responses.

Response variability differed substantially between correct and incorrect final answers (Figure 2A). When the majority-voted answer was correct, samples contained an average of 2.25 unique answers (SD = 0.77); when incorrect, samples contained 2.91 unique answers (SD = 0.65, Figure 2B). This difference was significant (Mann-Whitney U = 3,429,696, *p <* 10^−5^) with a large effect size (Cohen’s *d* = 0.90).

The distributional differences were notable (Kolmogorov-Smirnov statistic = 0.39, *p <* 10^−5^). Among correct responses, 15.3% showed complete agreement (1 unique answer) and 48.9% showed near-agreement (2 unique answers, Figure 2A). In contrast, only 0.7% of incorrect responses showed complete agreement, while 59.3% produced 3 unique answers and 16.0% produced 4 unique answers—indicating the model distributed probability mass across most available options.

These findings suggest that the agreement rate within P2 sampling functions as an implicit confidence measure. High agreement (1–2 unique answers) indicates reliable model knowledge and predicts correctness, while high variability (3–4 unique answers) signals uncertainty and predicts errors. This calibration property has practical implications: systems could flag low-agreement responses for human review or additional verification, potentially improving the safety of clinical decision support applications without requiring explicit uncertainty quantification methods.

### 2.5 PCCR iteration characteristics and answer stability

To characterize the computational mechanics of PCCR, we examined the number of continuation iterations required to reach each token budget threshold (Figure 3).

**Fig 3.**
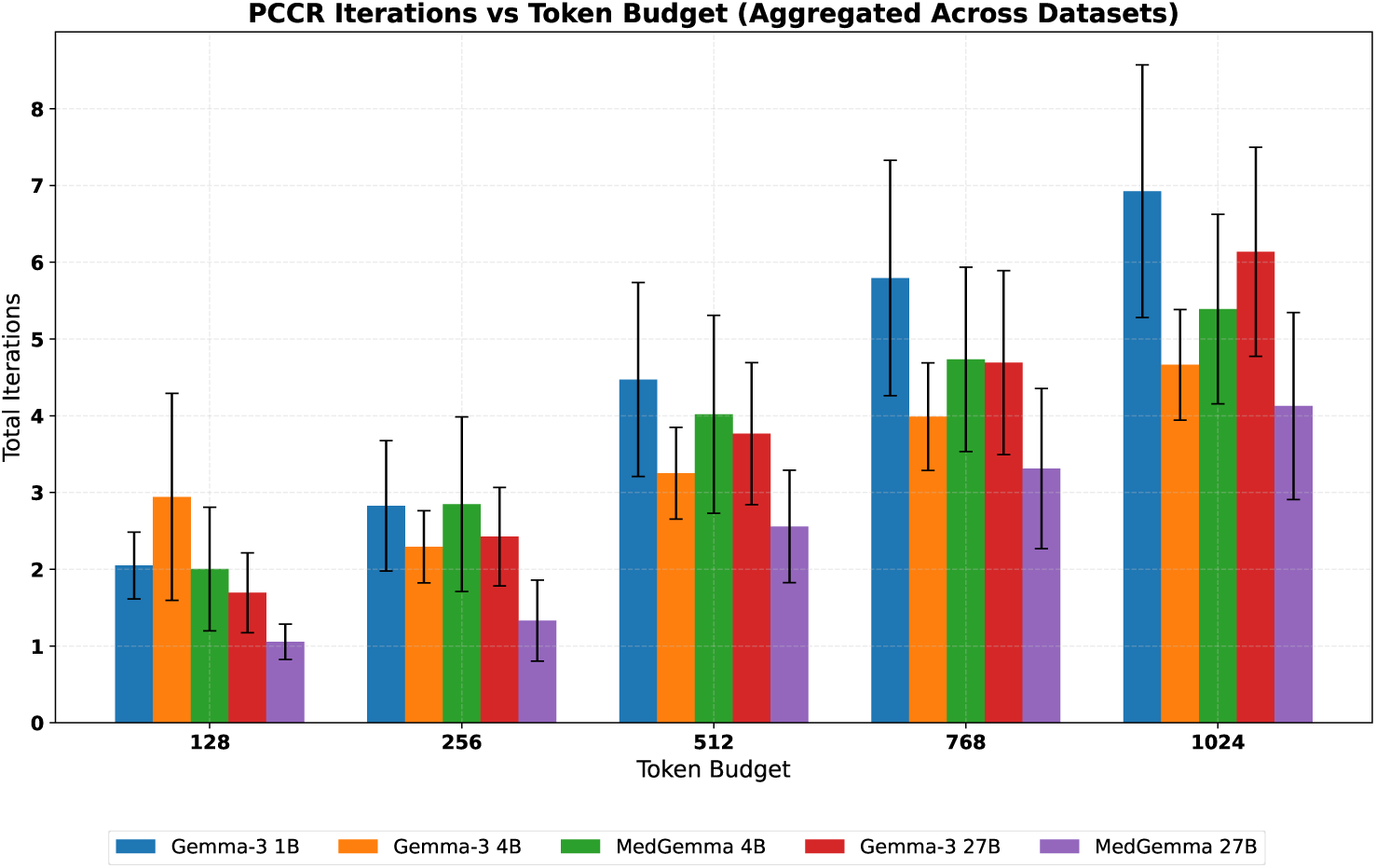
Number of continuation iterations required to reach PCCR token budgets. Mean ± SD number of prompting rounds (initial prompt plus continuations) required to meet each token budget threshold, aggregated across all datasets. Error bars indicate standard deviation. Smaller models (Gemma-3 1B) require more iterations to reach equivalent token budgets due to shorter per-response generation lengths, while larger models—particularly MedGemma 27B—produce longer reasoning traces per iteration, often meeting budget thresholds in fewer rounds. At PCCR-1024, Gemma-3 1B requires an average of 6.9 iterations compared to 4.1 for MedGemma 27B.

Iteration counts varied substantially by model size: smaller models produced shorter reasoning traces per prompt, requiring more continuation rounds to meet budget thresholds. At PCCR-1024, Gemma-3 1B required an average of 6.9 iterations (SD = 1.6), while MedGemma 27B required only 4.1 iterations (SD = 0.9). This pattern was consistent across budget levels, with the gap widening at higher thresholds.

Interestingly, Gemma-3 27B required more iterations than MedGemma 27B at most budget levels despite identical architectures, suggesting that medical fine-tuning encouraged more verbose reasoning traces.

We additionally investigated the stability of initial answers across PCCR continuation by examining answer flips—cases where a response that was correct after the initial prompt became incorrect after subsequent continuation rounds. This analysis illuminates the mechanism underlying the “overthinking” penalty observed with context. For MedGemma 27B on MIRAGE (no context), where PCCR-1024 substantially outperformed PCCR-128 (80.1% vs. 58.1%), only 1.8% of initially correct responses flipped to incorrect during continuation. In contrast, for MedGemma 4B on PubMedQA with oracle context, where PCCR-1024 substantially underperformed PCCR-128 (63.8% vs. 75.2%), 13.2% of initially correct responses flipped to incorrect—a 7.3-fold higher flip rate.

This disparity in flip rates provides mechanistic insight into when extended reasoning helps versus harms. Without context, models benefit from additional reasoning to access and organize parametric knowledge; initial responses are often uncertain and refinable. With high-quality context, initial responses are more likely to be correct context extractions; continued reasoning introduces opportunities to second-guess these correct answers, with each iteration carrying a probability of harmful revision. The substantially higher flip rate in the context condition (13.2% vs. 1.8%) quantifies this risk.

### 2.6 An Empirically Grounded Framework for Medical LLM System Design

Building on these empirical observations, we now formalize a framework for reasoning about the trade-offs between model scale, fine-tuning, test-time scaling, and context grounding when designing medical LLM systems.

#### 2.6.1 The accuracy model

Effective accuracy can be modeled as a function of model size (*S*), fine-tuning status (*F*), prompting strategy (*P*), reasoning budget (*b*), and context availability (*C*):

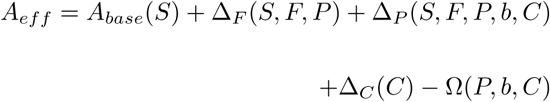

where:

- *A_base_*(*S*) is baseline accuracy as a function of model parameters
- Δ*_F_* (*S, F, P*) is the accuracy contribution from medical fine-tuning
- Δ*_P_* (*S, F, P, b, C*) is the accuracy contribution from prompting strategy and reasoning budget
- Δ*_C_*(*C*) is the accuracy contribution from context grounding
- Ω(*P, b, C*) is the overthinking penalty, which depends critically on prompting strategy

Now we describe each component of the accuracy model in detail.

### Baseline accuracy

From our empirical data, baseline accuracy (P0, no context) follows a logarithmic relationship with model scale:

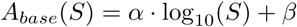

Fitting to our MIRAGE P0 results for base Gemma models yields *α* ≈ 14.4 and *β* ≈ 21.6, giving predicted values found in Table 8.

**Table 8.** Baseline accuracy model fit vs. observed from MIRAGE.

| Model Size | Predicted $A_{base}$ | Observed (Gemma-3) |
| --- | --- | --- |
| 1B | 35.0% | 34.6% |
| 4B | 43.3% | 44.1% |
| 27B | 54.2% | 55.3% |

### Fine-tuning contribution

The accuracy gain from medical fine-tuning depends on both model scale and prompting strategy, with an interaction term capturing the amplification effect observed with sophisticated prompting:

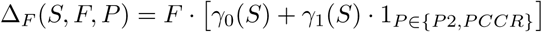

From our data at the 27B scale:

- *γ*_0_(27*B*) ≈ 4.6Δ% (fine-tuning benefit with simple prompting)
- *γ*_1_(27*B*) ≈ 9.5Δ% (additional benefit with sophisticated prompting)

This captures our finding that fine-tuning benefit increases from +4.6 Δ% (P0) to +13–16 Δ% (P2, PCCR) at the 27B scale on tasks without context.

### Prompting strategy contribution

The effect of prompting strategy varies substantially by model scale, context availability, and the specific strategy employed. Critically, our results reveal that P2 (self-consistency) and PCCR operate through distinct mechanisms with different context interactions:

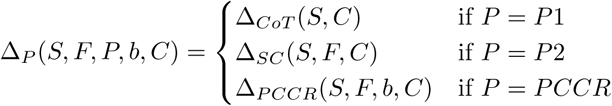

#### Chain-of-thought (P1)

The effect of single-pass chain-of-thought depends on model scale and context:

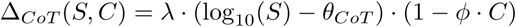

where *θ_CoT_* ≈ 0.9 (corresponding to an assumed ∼8B parameters) is the threshold below which CoT becomes harmful without context. The term (1 − *ϕ* · *C*) captures the observation that CoT benefits are attenuated when context is available—models can extract answers directly rather than reasoning from parametric knowledge.

#### Self-consistency (P2)

Self-consistency shows a qualitatively different pattern from extended sequential reasoning:

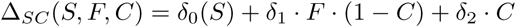

The key insight from our PubMedQA results is that P2 provides robust benefits with context across all model scales, including smaller models that show minimal P2 benefit without context. Table 9 presents the empirical P2 gains by context condition.

**Table 9.** Self-consistency (P2) accuracy gain over P0 by context availability.

| Model | P2 – P0 (no context) | P2 – P0 (with context) |
| --- | --- | --- |
| | MIRAGE ($\Delta\%$) | PubMedQA ($\Delta\%$) |
| MedGemma 27B | +17.0 | +0.5 |
| Gemma-3 27B | +5.9 | +1.1 |
| MedGemma 4B | +2.0 | −0.9 |
| Gemma-3 4B | −2.4 | −0.3 |
| Gemma-3 1B | −7.3 | −6.5 |

Without context, P2 benefits are concentrated in larger fine-tuned models (MedGemma 27B shows +17.0 Δ% on MIRAGE), while smaller and base models show minimal or negative effects. With context, the pattern reverses: base models show the largest P2 gains (+9.8 to +10.1Δ% for Gemma-3 4B and 27B), while already high-performing configurations show ceiling effects.

This pattern can be understood mechanistically. Without context, P2 benefits require the model to produce multiple valid reasoning paths that converge on correct answers—a capability that depends on model scale and domain knowledge. With context, even smaller models can extract correct answers from the provided evidence, but do so with variable reliability; P2 voting filters out erroneous extractions while preserving correct ones.

#### Prompt-chaining for continuous reflection (PCCR)

PCCR follows a saturating exponential relationship with reasoning budget, but critically, this relationship is modulated by context availability:

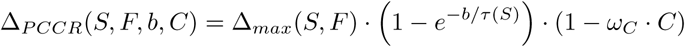

The term (1 − *ω_C_*· *C*) captures the reduced benefit of extended reasoning when context is available. From our data, *ω_C_*≈ 0.6–0.8, indicating that context reduces the value of extended reasoning by 60–80%.

Table 10 presents fitted parameters for the PCCR accuracy model from our MIRAGE data (no context condition). The model achieves good fit to the no-context data (mean absolute error = 0.8Δ%), validating the saturating exponential form for the no-context regime.

**Table 10.** Fitted parameters for the PCCR accuracy model on MIRAGE (no context).

| Model | $\Delta\%_{max}$ | $\tau$ (tokens) | $A_{P0}$ | $A_{PCCR-1024}$ pred. | $A_{PCCR-1024}$ obs. |
| --- | --- | --- | --- | --- | --- |
| MedGemma 27B | 22.0 | 350 | 59.9% | 79.8% | 80.1% |
| Gemma-3 27B | 12.5 | 400 | 55.3% | 66.5% | 66.8% |
| Gemma-3 4B | 7.0 | 250 | 44.1% | 50.2% | 49.1% |
| MedGemma 4B | 4.0 | 300 | 47.2% | 50.6% | 50.6% |
| Gemma-3 1B | 12.0 | 500 | 34.6% | 44.9% | 46.2% |

### Context contribution

Context grounding provides a substantial additive benefit:

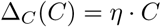

From our PubMedQA data comparing no-context to oracle-context conditions at P0, *η* ranges from approximately 18 to 32Δ% depending on model. For planning purposes, we estimate *η* ≈ 25Δ% for oracle-quality retrieval.

### Overthinking penalty

A critical finding from our experiments is that the overthinking penalty depends strongly on prompting strategy. We model this as:

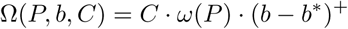

where (*x*)^+^ = max(0*, x*) and the penalty rate *ω*(*P*) varies by prompting strategy:

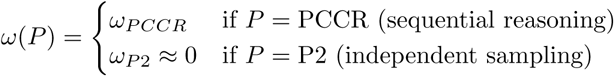

Our PubMedQA results demonstrate that P2 is largely immune to the overthinking penalty that affects PCCR. Table 11 compares the two strategies with oracle context. P2 matches or exceeds the best non-PCCR configuration for 4 of 5 models, while PCCR-1024 underperforms for 4 of 5 models. The average P2 deviation from best is +2.1Δ% (beneficial), while the average PCCR-1024 deviation is −4.5Δ% (harmful). This divergence reflects the fundamental mechanistic difference: P2 samples independent reasoning chains and aggregates via voting, providing robustness against overthinking, while PCCR accumulates reasoning within a single chain, allowing errors to compound.

**Table 11.** Overthinking penalty comparison: P2 (self-consistency) versus PCCR-1024 with oracle context on PubMedQA.

| Model | Best non-PCCR | P2 | PCCR-1024 | P2 vs Best | PCCR-1024 vs. Best |
| --- | --- | --- | --- | --- | --- |
| MedGemma 27B | 77.4% (P1) | 75.2% | 70.1% | $-2.2\Delta\%$ | $-7.3\Delta\%$ |
| Gemma-3 27B | 68.4% (P1) | 70.1% | 61.3% | $+1.7\Delta\%$ | $-7.1\Delta\%$ |
| MedGemma 4B | 73.1% (P0) | 74.3% | 63.8% | $+1.2\Delta\%$ | $-9.3\Delta\%$ |
| Gemma-3 4B | 64.7% (P1) | 70.1% | 65.3% | $+5.4\Delta\%$ | $+0.6\Delta\%$ |
| Gemma-3 1B | 58.6% (P1) | 63.1% | 59.0% | $+4.5\Delta\%$ | $+0.4\Delta\%$ |

For PCCR specifically, we estimate the overthinking penalty rate from PubMedQA with context in Table 12.

**Table 12.** Overthinking penalty parameters for PCCR with oracle context.

| Model | $b^*$ (optimal tokens) | $\omega_{PCCR}$ ($\Delta\%$ per 100 tokens) |
| --- | --- | --- |
| MedGemma 27B | $\sim 0$ (P1 optimal) | 0.8 |
| Gemma-3 27B | $\sim 256$ | 1.2 |
| MedGemma 4B | $\sim 256$ | 1.6 |
| Gemma-3 4B | $\sim 128$ | 1.3 |
| Gemma-3 1B | $\sim 128$ | 1.0 |

These values indicate that for PCCR, each additional 100 tokens of reasoning beyond the optimum costs approximately 0.8–1.6Δ% of accuracy when good context is available.

#### 2.6.2 The cost model

Total cost per query combines inference cost and retrieval cost:

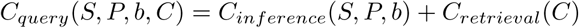

### Inference cost

Inference cost depends on input and output token counts, which vary by prompting strategy:

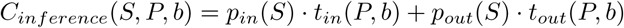

Table 13 presents the token counts for each prompting strategy.

**Table 13.** Token consumption by prompting strategy.

| Strategy | Input Tokens | Output Tokens | Total Relative Cost |
| --- | --- | --- | --- |
| P0 | $t_q$ | $t_a \approx 20$ | $1\times$ |
| P1 | $t_q$ | $t_a + t_{cot} \approx 200$ | $\sim 3\times$ |
| P2 | $5 \cdot t_q$ | $5 \cdot (t_a + t_{cot}) \approx 1000$ | $\sim 15\times$ |
| PCCR-$b$ | $t_q + k \cdot b$ | $b + t_a$ | $\sim (2-10)\times$ depending on $b$ |

For PCCR, the number of continuation rounds *k* depends on the natural reasoning length per prompt *b_step_*: where *b_step_* ≈ 100–200 tokens based on our observations.

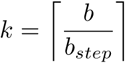

Our results reveal that P2 and PCCR have similar token costs at high reasoning budgets (P2≃1000 tokens, PCCR-1024≃1024 tokens), but dramatically different effectiveness when context is available. With oracle context on PubMedQA, P2 achieves 63.1-75.2% accuracy while PCCR-1024 achieves 59-70.1% accuracy across models. At equivalent cost, P2 provides higher accuracy on average when context is available, making the dominant strategy for RAG-based applications.

Table 14 presents representative API pricing for the Gemma model family.

**Table 14.** Representative API pricing for Gemma models (USD per million tokens).

| Model | Input Price | Output Price | Source |
| --- | --- | --- | --- |
| Gemma-3 1B | \$0.01* | \$0.01* | *Generally free-tier |
| Gemma-3 4B | \$0.02 | \$0.04 | DeepInfra |
| Gemma-3 27B | \$0.10 | \$0.20 | DeepInfra |

### Retrieval cost

When using RAG, additional costs include embedding computation and context tokens:

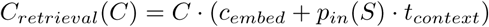

where *c_embed_* is the cost of embedding the query and *t_context_* is the number of retrieved context tokens (typically 500–3000). We discuss the additional costs of RAG and fine-tuning in more detail below in Section 2.7.3 as well as Appendix E and F.

### The latency model

Total latency comprises retrieval latency (if applicable), prefill latency, and decode latency:

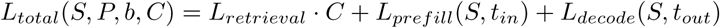

Decode latency depends on model throughput *τ* (*S*) (tokens per second):

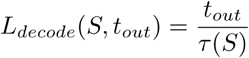

### P2 latency advantage with parallelization

A key practical consideration is that P2’s five samples can be generated in parallel, while PCCR’s continuation rounds are inherently sequential. With parallel execution:

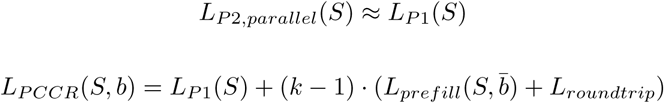

This means P2 achieves its accuracy benefits with latency comparable to single sample P1, while PCCR incurs multiplicative latency from sequential continuation rounds. For latency-sensitive applications with high-quality context, P2 is strongly preferred. Table 15 presents representative throughput values (estimation methodology in Appendix C).

**Table 15.** Estimated theoretical inference throughput (tokens/second) based on memory bandwidth utilization (Roofline model). Estimates assume 0.65 bytes/param for INT4 and 2.0 bytes/param for FP16.

| Model | RTX 4090 (INT4) | A100 (FP16) | Consumer CPU |
| --- | --- | --- | --- |
| Gemma-3 1B | ~1,400* | ~880* | ~85 |
| Gemma-3 4B | ~370 | ~230 | ~22 |
| Gemma-3 27B | ~57 | ~35 | ~3 |

\*Theoretical bandwidth limit. Real-world small-model performance is often latency-bound (approx. 250–300 t/s) unless specialized kernels are used.

### 2.7 Solving the optimization problem

We now demonstrate that the optimization problem defined by our framework is tractable and can be solved analytically or numerically to yield optimal configurations. We first formalize the complete objective and constraints, then solve for two representative scenarios that describe a fundamental bifurcation of optimal strategy. We describe clear heuristic constraints identified by our framework and provide recommendations for configuration selection. We also provide further examples of solving the optimization problem in Appendix G.

#### 2.7.1 Problem Formulation

The system designer seeks to minimize cost subject to accuracy (*A_min_*) and latency constraints (*L_max_*):

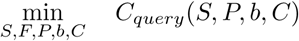

subject to:

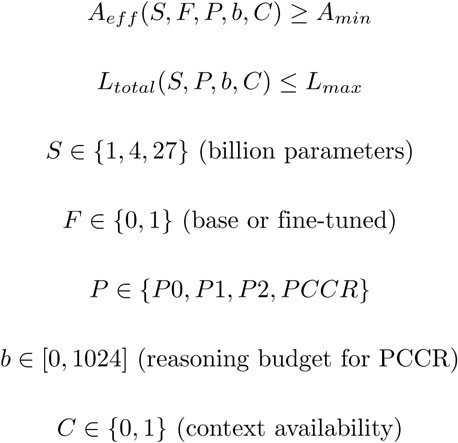

#### 2.7.2 Accuracy look-up table

Given the discrete nature of our experimental conditions and the complex interactions observed, we construct a look up table relating system configurations to accuracy, *A*^^^(*S, F, P, b, C*). Table 16 presents the complete accuracy surface.

**Table 16.**
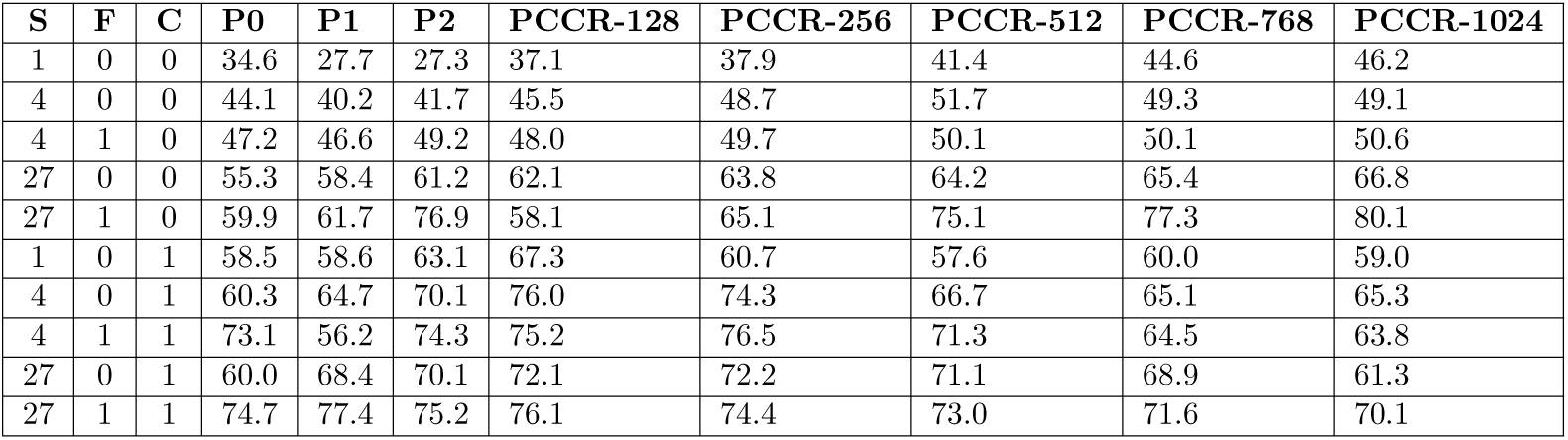
Accuracy values (%) used for optimization. Interpolated values for PCCR budgets between measured points use linear interpolation.

| S | F | C | P0 | P1 | P2 | PCCR-128 | PCCR-256 | PCCR-512 | PCCR-768 | PCCR-1024 |
| --- | --- | --- | --- | --- | --- | --- | --- | --- | --- | --- |
| 1 | 0 | 0 | 34.6 | 27.7 | 27.3 | 37.1 | 37.9 | 41.4 | 44.6 | 46.2 |
| 4 | 0 | 0 | 44.1 | 40.2 | 41.7 | 45.5 | 48.7 | 51.7 | 49.3 | 49.1 |
| 4 | 1 | 0 | 47.2 | 46.6 | 49.2 | 48.0 | 49.7 | 50.1 | 50.1 | 50.6 |
| 27 | 0 | 0 | 55.3 | 58.4 | 61.2 | 62.1 | 63.8 | 64.2 | 65.4 | 66.8 |
| 27 | 1 | 0 | 59.9 | 61.7 | 76.9 | 58.1 | 65.1 | 75.1 | 77.3 | 80.1 |
| 1 | 0 | 1 | 58.5 | 58.6 | 63.1 | 67.3 | 60.7 | 57.6 | 60.0 | 59.0 |
| 4 | 0 | 1 | 60.3 | 64.7 | 70.1 | 76.0 | 74.3 | 66.7 | 65.1 | 65.3 |
| 4 | 1 | 1 | 73.1 | 56.2 | 74.3 | 75.2 | 76.5 | 71.3 | 64.5 | 63.8 |
| 27 | 0 | 1 | 60.0 | 68.4 | 70.1 | 72.1 | 72.2 | 71.1 | 68.9 | 61.3 |
| 27 | 1 | 1 | 74.7 | 77.4 | 75.2 | 76.1 | 74.4 | 73.0 | 71.6 | 70.1 |

#### 2.7.3 Cost model

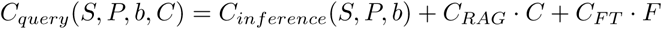

where inference cost depends on token counts:

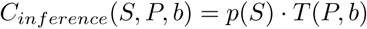

Using blended pricing *p*(*S*) (average of input/output at typical ratios) and token multipliers *T* (*P, b*):

| S | $p(S)$ (\$/1M) |
| --- | --- |
| 1 | \$0.01 |
| 4 | \$0.03 |
| 27 | \$0.10 |

| P | $T(P, b)$ (tokens) |
| --- | --- |
| P0 | 120 |
| P1 | 300 |
| P2 | 1,500 |
| PCCR-$b$ | $300 + 1.5b$ |

RAG and model fine-tuning incur different upfront and recurring costs over different time spans. To account for them at the level of the query, we must amortize them by the expected query volume over a fixed duration. For convenience and common accounting practices, we select this duration to be 1 year and use reasonable assumptions to pick a reference query volume of 20 million queries per year to amortize upfront and recurring costs (justified in Appendix D).

RAG overhead: *C_RAG_* = $0.0001 per query based on reference query volume (embedding + retrieval infrastructure amortized cost, estimation in Appendix E).

Fine-tuning overhead: *C_F T_* = $0.0013 per query based on reference query volume (data collection + training compute amortized cost, estimation in Appendix F).

The RAG and fine-tuning costs are amortized using the value for the query volume. Increases or reductions in the query volume will result in corresponding linear changes to reference values for *C_RAG_* and *C_F T_* that were used in this analysis. However, the relative difference between *C_RAG_* and *C_F T_* would be maintained. At very high query volumes, *C_RAG_* may be comparable to the cost of token generation, *C_inference_*, but *C_F T_* may remain high.

#### 2.7.4 Latency model

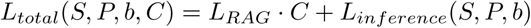

where:

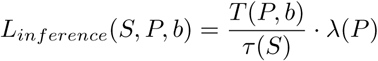

We limit token generation limits to 300 tokens/sec due to realistic latency and bandwidth limitations. We approximate RAG latency to be *L_RAG_* = 0.3 seconds

| **S** | $\tau(S)$ (tokens/sec) |
| --- | --- |
| 1 | 300 |
| 4 | 200 |
| 27 | 35 |

| **P** | $\lambda(P)$ (parallelization factor) |
| --- | --- |
| P0 | 1.0 |
| P1 | 1.0 |
| P2 | 1.0 (parallel sampling) |
| PCCR | $k$ (sequential rounds, $k = \lceil b/150 \rceil$) |

#### 2.7.5 Analytical solution approach

Given the discrete nature of model size, fine-tuning status, prompting strategy, and context availability, the optimization reduces to enumeration over a finite set of configurations with optimization over *b* for PCCR strategies.

For each discrete configuration (*S, F, P, C*) with *P* ≠ *PCCR*:

1. Compute *A_eff_* from Table 16
2. Compute *C_query_* and *L_total_* from cost/latency models
3. Check feasibility: *A_eff_* ≥ *A_min_* and *L_total_* ≤ *L_max_*

For PCCR configurations, we additionally optimize over *b*:

1. Find minimum *b*^∗^ such that *A_eff_* (*S, F, PCCR, b*^∗^*, C*) ≥ *A_min_* (if exists)
2. Check latency feasibility at *b*^∗^
3. Compute cost at *b*^∗^

The global optimum is the feasible configuration with minimum cost. In Figure 4, we visualize the accuracy-cost trade-off of each model-prompt configuration when considering model size, context-availability, and fine-tuning. The figure clearly shows the effect on accuracy from the availability of context vs. no-context (red vs. blue) and the effect on cost from fine-tuned vs. generic models (far-right vs. far-left).

**Fig 4.**
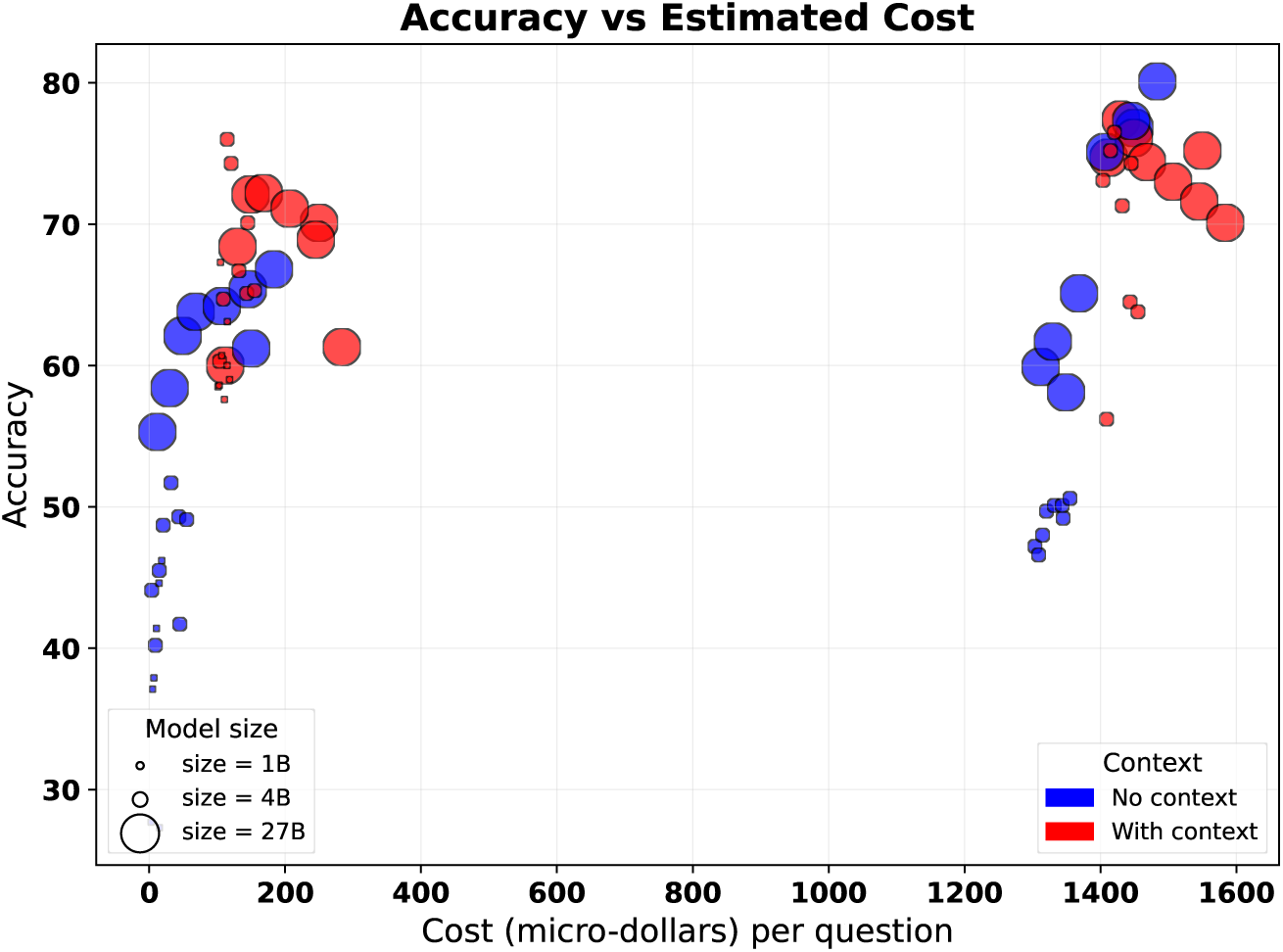
Accuracy vs. estimated cost per question in $1×10^−6^, or *µ*$, based on the empirically grounded framework described in Section 2.6 for LLMs of different sizes (size of the marker) with context (red) or with no context (blue). Each point is a model-prompt configuration but the prompt-style is not represented here. Medically fine-tuned models are on the far right with 7-15× the cost of generic models. Detailed assumptions for estimated RAG and fine-tuning costs per question can be found in Appendix E & F.

#### 2.7.6 Example 1: High-accuracy RAG application

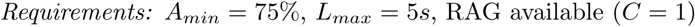

We enumerate candidate configurations from Table 16 with *C* = 1 and achieving ≥ 75% to populate Table 17. The optimal solution configuration is (4, 0, PCCR-128, 1) or Gemma-3 4B + PCCR-128 + RAG at 114.76*µ*$ per query and a latency of 2.76 seconds.

**Table 17.** Candidate configurations for Example 1: High-accuracy RAG application that meet the accuracy requirement. Values shown in micro-dollars (*µ*$ = 10^−6^ USD).

| Config | Accuracy | Cost | Latency | Feasible? |
| --- | --- | --- | --- | --- |
| (27, 1, P1, 1) | 77.4% | 1412$\mu\$$ | 8.9s | No (latency) |
| (27, 1, P2, 1) | 75.2% | 1550$\mu\$$ | 8.9s | No (latency) |
| (27, 1, PCCR-128, 1) | 76.1% | 1449.2$\mu\$$ | 14.34s | No (latency) |
| (4, 1, PCCR-256, 1) | 76.5% | 1420$\mu\$$ | 5.0s | Yes |
| (4, 1, PCCR-128, 1) | 75.2% | 1414.76$\mu\$$ | 2.76s | Yes |
| (4, 0, PCCR-128, 1) | 76.0% | 114.76$\mu\$$ | 2.76s | Yes |

Note that fine-tuning incurs a notable cost when high-quality context can provide answers to questions, with fine-tuned candidate configurations being 10× more expensive with no substantial gain in accuracy.

#### 2.7.7 Example 2: No-context, high-accuracy application

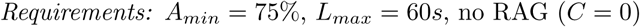

We enumerate candidate configurations from Table 16 with *C* = 0 and achieving ≥ 75% to populate Table 18. The optimal solution configuration is (27, 1, PCCR-512, 0) or MedGemma 27B + PCCR-512 at 1406.8*µ*$ per query and a latency of 30.51 seconds. Without appropriate context that contains answers or questions that require reasoning, the system designer is forced to use a fine-tuned model with higher costs. Notably, for *<* 100*µ*$ more per query, the system designer could choose a configuration with a higher accuracy, (27, 1, PCCR-1024, 0), that still fits the latency requirement.

**Table 18.** Candidate configurations for Example 2: No-context, high-accuracy application that meet the accuracy requirements. Values shown in micro-dollars (*µ*$ = 10^−6^ USD).

| Config | Accuracy | Cost | Latency | Feasible? |
| --- | --- | --- | --- | --- |
| (27, 1, P2, 0) | 76.9% | $\$1450.0\mu$ | 8.57s | **Yes** |
| (27, 1, PCCR-512, 0) | 75.1% | $\$1406.8\mu$ | 30.51s | **Yes** |
| (27, 1, PCCR-768, 0) | 77.3% | $\$1445.2\mu$ | 41.49s | **Yes** |
| (27, 1, PCCR-1024, 0) | 80.1% | $\$1483.6\mu$ | 52.46s | **Yes** |

##### Strategy Selection Based on Context Availability

As highlighted by the two examples, our results suggest a fundamental bifurcation in optimal strategy based on context quality, informing general heuristics for system design:

When high-quality context is available (RAG with reliable retrieval):

1. Generally, prefer shorter reasoning traces (P1 or PCCR-128) over extended reasoning budgets that are counterproductive—limit *b* ≤ *b*^∗^(*S*).
2. Consider P2 for confidence requirements but this will also incur modestly higher costs due to token counts.
3. Small models (1B / 4B) can match large models (27B) with appropriate prompting
4. Fine-tuning benefit is reduced (context provides domain knowledge)

When context is unavailable or unreliable:

1. Prefer PCCR (extended reasoning) for maximum accuracy
2. Fine-tuning provides substantial benefit, especially with sophisticated prompting
3. Larger models significantly outperform smaller ones
4. P2 benefit is concentrated in large, fine-tuned models

In Table 19, we provide recommended configurations based on objectives and rationale that map to the heuristics provided above and the costs discussed previously.

**Table 19.** Recommended prompting strategy by context availability and primary objective.

| Context | Objective | Recommended Strategy | Rationale |
| --- | --- | --- | --- |
| High-quality | Max accuracy | P2 or PCCR-128 | Reduce overthinking; voting filters errors |
|  | Min latency | P0, P1, or PCCR-128 | Direct extraction sufficient |
|  | Min cost | P0 | Minimal tokens; context provides knowledge |
| Low/none | Max accuracy | PCCR-1024 (MedGemma 27B) | Extended reasoning helps access parametric knowledge |
|  | Balanced | P2 (MedGemma 27B) | Strong gains with moderate cost |
|  | Min cost | P0 (largest affordable model) | Reasoning overhead not cost-effective at small scale |

#### 2.7.8 Equivalence points

Using our framework, we can identify configurations where smaller or simpler systems achieve equivalent accuracy to larger ones. Table 20 presents key equivalence points derived from our empirical data. The equivalence points with RAG are particularly striking: Gemma-3 4B with PCCR-128 and context achieves 76.0% accuracy, compared to MedGemma 27B with P1 and context at 77.4%—within confidence intervals at approximately 10% of the cost. The effect of fine-tuning on estimated cost is evident in the first row of Table 20 where MedGemma 27B with P0 is comparable in accuracy of Gemma-3 27B with PCCR-256 but the Gemma-3 27B model is 2% of the cost-per-query.

**Table 20.** Equivalence points: configurations achieving comparable accuracy at different cost/latency profiles.

| Reference Configuration | Accuracy | Equivalent Configuration | Accuracy | Cost Ratio | Context Required |
| --- | --- | --- | --- | --- | --- |
| MedGemma 27B P0 (MIRAGE) | 59.9% | Gemma-3 27B PCCR-256 | 63.8% | 0.02× | No |
| MedGemma 27B P2 (MIRAGE) | 76.9% | MedGemma 27B PCCR-512 | 75.1% | 0.9× | No |
| MedGemma 27B P1 + RAG | 77.4% | Gemma-3 4B P2 + RAG | 70.1% | 0.10× | Yes |
| MedGemma 27B P1 + RAG | 77.4% | MedGemma 4B PCCR-256 + RAG | 76.5% | 0.99× | Yes |
| MedGemma 27B P1 + RAG | 77.4% | Gemma-3 4B PCCR-128 + RAG | 76.0% | 0.08× | Yes |
| Gemma-3 27B P0 + RAG | 60.0% | Gemma-3 1B PCCR-128 + RAG | 67.3% | 0.92× | Yes |

Even Gemma-3 1B is able to perform comparably to Gemma-3 27B under certain configurations with high-quality context. This suggests that for RAG-based applications, investment in retrieval quality may yield better returns than investment in model scale.

## 3 Discussion

This study systematically evaluated the trade-offs between model scale, medical fine-tuning, test-time scaling, and context grounding for medical question-answering using the Gemma and MedGemma model families. Our findings reveal complex interactions between these optimization dimensions that have practical implications for clinical AI system design.

### 3.1 Summary of key empirical findings

#### Chain-of-thought prompting harms smaller models

Contrary to the widespread assumption that explicit reasoning improves performance, we found that P1 prompting decreased accuracy for smaller models. On MIRAGE, Gemma-3 1B dropped 6.9 percentage points and Gemma-3 4B dropped 3.9 percentage points when prompted to reason step-by-step (Table 2). This pattern replicated across datasets, with Gemma-3 1B showing a 22.2 percentage point drop on PubMedQA without context (Table 4). These results suggest that chain-of-thought reasoning requires sufficient model capacity to execute coherently; smaller models appear to “talk themselves out of” correct initial responses.

#### Medical fine-tuning benefits are amplified by sophisticated prompting

The accuracy advantage of MedGemma over base Gemma models grew substantially when combined with self-consistency or extended reasoning. On MIRAGE, the fine-tuning benefit at the 27B scale increased from +4.6 percentage points with direct answering (P0) to +15.7 percentage points with self-consistency (P2)—a 3.4-fold amplification (Table 2). This interaction effect indicates that evaluations using only simple prompting strategies substantially underestimate the value of medical fine-tuning.

#### Extended sequential reasoning (PCCR) enables continuous accuracy scaling without context

Our prompt-chaining method demonstrated monotonically increasing accuracy with reasoning budget when context was unavailable. MedGemma 27B improved from 59.9% (P0) to 80.1% (PCCR-1024) on MIRAGE—a gain of 20.2 percentage points (Table 2). Even Gemma-3 1B showed substantial improvement, increasing from 34.6% to 46.2% with extended reasoning. These gains demonstrate that inference-time computation can partially substitute for model scale and domain-specific training.

#### Context grounding provides substantial but interaction-dependent benefits

Oracle context improved accuracy by 18–32 percentage points across models on PubMedQA, demonstrating the value of retrieval-augmented generation. However, the benefit of context interacted strongly with prompting strategy—a finding with important implications for system design.

#### Extended reasoning with context causes “overthinking.”

When high-quality context was available, PCCR accuracy degraded with increasing reasoning budget. MedGemma 27B peaked at 77.4% with P1 and declined to 70.1% at PCCR-1024—a 7.3 percentage point loss from extended reasoning. This pattern was consistent across all models, with overthinking penalties ranging from 7 to 13 percentage points. The effect suggests that models second-guess correct context-derived answers when forced to reason extensively.

#### Self-consistency (P2) is robust to overthinking

In contrast to PCCR, self-consistency voting maintained or improved accuracy with context across all model scales. On PubMedQA with oracle context, P2 achieved the highest or near-highest accuracy for most models, with base Gemma models showing +4.6 to +10.1 percentage point gains over P0. Notably, P2 recovered entirely from MedGemma 4B’s anomalous P1 degradation (56.2%→74.3%). This robustness likely stems from P2’s independent sampling mechanism: majority voting filters out erroneous reasoning chains while preserving correct context extractions, rather than accumulating errors as in sequential reasoning.

#### Small models with context can match larger models

With oracle context and appropriate prompting, 4B models achieved accuracy comparable to 27B models. Gemma-3 4B with PCCR-128 reached 76.0%, approaching MedGemma 27B with P1 at 77.4%. Even Gemma-3 1B with PCCR-128 (67.3%) exceeded Gemma-3 27B with P0 (60.0%). These equivalence points suggest that investment in retrieval infrastructure may yield better accuracy-per-dollar than investment in larger and larger models for many clinical applications. Critically, test-time scaling methods without context failed to close the gap between 4B models and 27B models in our experiments. However, the 1B model under certain configurations performed as well as the 4B model in every dataset we tested, suggesting that test-time scaling methods are fungible with increases in model size up to a certain limit. From our experiments, we approximate this limit to be between 4 × −7× based on the size ratios of 4B:1B and 27B:4B models.

#### Optimal strategy depends critically on context availability

Our results reveal a fundamental bifurcation: without context, extended reasoning (PCCR) and fine-tuning provide the greatest benefits; with high-quality context, simpler prompting (P0, P1) or self-consistency (P2) is preferred, and extended reasoning becomes counterproductive. This finding implies that adaptive systems should dynamically adjust prompting strategy based on retrieval confidence.

### 3.2 Revisiting the scaling paradigm

The dominant paradigm in medical AI development has emphasized model scale as the primary lever for achieving clinical competence. Our results suggest a more nuanced picture: scale remains important, but its benefits can be substantially replicated through alternative strategies. With high-quality context, the accuracy of 4B models with short extended reasoning are comparable with that of fine-tuned 27B models—at approximately one-tenth the computational cost. Similarly, without context on the MIRAGE datset, MedGemma 27B with extended reasoning (PCCR-1024) reached 80.1% which is higher than the 73.4% accuracy reported for GPT-4 without context and matched the performance of GPT-4 with access to extensive context [21].

These equivalence points have practical significance for healthcare organizations facing deployment constraints. Institutions requiring on-premise deployment for data privacy may be unable to host large parameter models (32B+) on available hardware or be unwilling to send sensitive data outside of their network. Our results indicate that smaller models, when combined with effective retrieval and appropriate prompting, can achieve useful accuracy levels. The critical insight is that scale, fine-tuning, retrieval, and inference-time computation are partially fungible—deficits in one dimension can be compensated by investments in others.

However, this fungibility has limits. For the most demanding accuracy requirements (*>* 77% on our benchmarks without context), MedGemma 27B with sophisticated prompting remained necessary. The choice of optimization strategy should therefore be guided by specific accuracy requirements, not assumptions about universal best practices.

### 3.3 Towards adaptive medical AI systems

Our empirically grounded framework formalizes the trade-offs revealed by our experiments. The framework’s practical value lies in enabling systematic configuration selection. Rather than defaulting to the largest available model or universally applying chain-of-thought prompting, practitioners can navigate the trade-off space based on specific requirements. Our recommendations provide explicit guidance: when high-quality retrieval is available, prefer self-consistency or simple prompting over extended reasoning; when retrieval is unavailable, invest in extended reasoning and larger fine-tuned models; when latency is critical, accept accuracy trade-offs from simpler configurations.

The equivalence points identified by our framework are particularly actionable and insightful. The finding that Gemma-3 1B and 4B with context achieve accuracies comparable to MedGemma 27B at one-tenth the cost suggests that many clinical applications may be over-provisioned. Conversely, applications currently using small models with simple prompting may achieve substantial accuracy gains by adding retrieval infrastructure or self-consistency voting. Critically, given cost and latency considerations, institutions considering deploying LLM-based assistants should invest in high-quality retrieval systems.

### 3.4 Clinical implications

While our evaluation focused on benchmark performance, several findings have clinical relevance. The overthinking phenomenon suggests that clinical decision support systems using RAG should be designed to trust high-quality retrieved evidence rather than extensively deliberating over it.

The robustness of self-consistency voting with context suggests a practical strategy for clinical applications: when uncertainty is a concern, sample multiple responses, potentially from multiple different retrievals, and aggregate rather than generating a single extended reasoning chain. This approach provides calibrated confidence (via agreement rates) while avoiding the overthinking trap and can be used to inform the confidence in the LLM response [28].

The chain-of-thought findings caution against deploying smaller models with reasoning prompts in clinical settings. If resource constraints necessitate smaller models (like mobile deployments), direct answering or externally structured reasoning (as in our PCCR approach) may be safer than prompting the model to reason in ways that cause issues.

Finally, the equivalence points suggest opportunities for democratizing medical AI. Smaller institutions unable to deploy large models may achieve comparable accuracy through investment in retrieval infrastructure—a more accessible path to clinical AI capabilities than acquiring high-end computational resources. Importantly, improving retrieval quality maybe more cost-effective than extended reasoning, using larger models, or fine-tuning models.

### 3.5 Limitations

Our evaluation used multiple-choice question-answering benchmarks, which, while standard in the field, do not fully capture the complexity of clinical reasoning. Real clinical decisions involve ambiguity, incomplete information, longitudinal context, and consequences that benchmarks cannot represent. Performance on MedQA-style questions may not translate directly to clinical utility, and our findings should be validated in more realistic clinical evaluation settings before informing deployment decisions.

Our PubMedQA experiments with context used oracle retrieval—the exact passage containing the answer. Real RAG systems achieve lower retrieval precision, and the interaction between retrieval quality and prompting strategy may differ when context is noisy or partially relevant. The overthinking phenomenon may be attenuated or reversed when retrieved context is unreliable and genuinely benefits from critical evaluation.

Future work should evaluate these interactions across a range of retrieval quality levels. No model configuration achieved the 65% passing threshold on the medical licensing examination, and all models performed below the median physician score of 70–80% [23].

The best model performances fell within the lowest quartile of physician performance (40–65%). This contrasts with reported results on established benchmarks like MedQA, where MedGemma 27B achieves 87.7% [15] and our own results on the MIRAGE dataset. This may be because the medical licensing examination questions were released recently, from a country other than the US, and, therefore, are less likely to be similar to the model training data, representing a more rigorous test of generalization.

Despite suboptimal absolute performance on the medical exam dataset, our findings remain important. The relative patterns we observed were consistent across all three datasets including the medical examination. These relationships hold regardless of absolute accuracy level. Our results may also provide a realistic assessment of open-weight model capabilities on genuinely novel content, which may better reflect real-world deployment scenarios than potentially contaminated benchmarks.

We emphasize that current open-weight models do not achieve physician-level performance on novel medical content and *should not be deployed as autonomous clinical decision-makers*. Understanding how to maximize their capabilities within current limitations remains essential for responsible medical AI development.

Our experiments focused on the Gemma and MedGemma model families. While these models are representative of current open-weight LLMs, our specific findings—including the chain-of-thought threshold, overthinking penalty rates, and equivalence points—may not generalize to other architectures. The MedGemma models incorporate specific training procedures and data that may produce different prompting response patterns than other medical LLMs. Replication across model families (e.g., Llama, Mistral, Qwen) would strengthen the generalizability of our conclusions.

We evaluated four prompting strategies, but the space of possible approaches is vast. Other techniques—including tree-of-thought, self-reflection, debate, and tool-augmented reasoning—may show different patterns [30–35]. However, our prompt-chaining for continuous reflection (PCCR) method shares conceptual foundations with several established approaches for eliciting improved reasoning from language models. While PCCR does not explicitly maintain a tree structure like tree-of-thought, the instruction to reflect on prior reasoning encourages implicit backtracking and branch exploration within the accumulated reasoning trace. PCCR also implements a lightweight form of self-reflection through its continuation prompt, which instructs the model to “verify your logic, evaluate differential diagnoses you may have overlooked, and ensure your reasoning is complete.” The instruction to continue reasoning—rather than to simply confirm or extend the existing answer—creates implicit pressure to consider opposing viewpoints, as in debate. Our approach offers practical advantages: it requires no specialized infrastructure (unlike multi-agent debate), no modifications to the inference process (unlike ToT’s tree search), and operates through natural language instructions compatible with any API (unlike methods requiring direct token manipulation). The trade-off is less explicit control over the deliberation structure. However, our empirical results demonstrate that even this simple operationalization of extended deliberation yields substantial accuracy improvements. Regardless, our PCCR method represents one approach to enforcing reasoning budgets; alternative implementations might yield different accuracy-budget relationships.

Moreover, PCCR iteration counts may have implications for network latency (Figure 3). Because PCCR continuation rounds are sequential (each round requires the previous response as input), total network latency scales multiplicatively with iteration count. Models with more iterations will require more round-trips to reach equivalent reasoning budgets. For systems where LLMs are being accessed over network connections that are not localized, the latency estimates for PCCR may be higher than what was modeled here.

Our evaluation was conducted primarily on English-language benchmarks. Medical AI systems deployed globally must handle diverse languages, and prompting strategy effects may vary across linguistic contexts.

Our framework treats accuracy, cost, and latency as static properties of configurations. In practice, these may vary with query characteristics, user context, and system load. A fully adaptive system would dynamically select configurations based on runtime signals, an approach our current framework supports conceptually but does not empirically validate. Critically, we use representative values for query-volume, infrastructure costs, and recurring costs that effect the cost-per-query estimates. These would need to be updated with real values given a use-case. While we do not expect changes in the general patterns in our framework, the specifics of query-volume, vector database costs, GPU-server costs, and cost-per-data-sample for fine-tuning can change the cost-per-query estimates presented here.

Our PCCR experiments enforced minimum token budgets through prompt continuation, but the relationship between token count and reasoning quality is imperfect. Models may generate padding or repetitive content to reach budgets without proportionally improving reasoning. More sophisticated methods for ensuring reasoning quality at target compute levels may yield different accuracy-budget curves.

### 3.6 Future work

Our results demonstrate that optimal prompting depends on context availability, but current systems typically apply fixed strategies. Developing methods that dynamically select prompting approaches based on retrieval confidence, query complexity, or model uncertainty could capture benefits across regimes. Initial work might focus on simple classifiers that route queries to different prompting pipelines based on retrieval scores.

We evaluated only oracle and no-context conditions. Systematic evaluation across retrieval quality levels—from perfect to noisy to adversarial—would characterize how prompting strategy recommendations vary with RAG system performance [36].

Understanding the threshold at which extended reasoning becomes beneficial for evaluating unreliable context is particularly important for practical deployment.

Replicating our experiments across diverse model families would establish which findings are architecture-specific versus general properties of LLM medical reasoning. Of particular interest is whether the chain-of-thought threshold and overthinking penalty rates are consistent across models or reflect idiosyncrasies of the Gemma architecture and training. Moreover, testing the recent “reasoning” models and understanding how medical reasoning is similar or different from mathematical or logical reasoning could help make significant advances in the applications of LLMs to clinical problems [37].

Moving beyond benchmarks to clinical evaluation settings—including physician assessment of response quality, simulation-based testing, and retrospective analysis on clinical data—would establish the clinical validity of our framework’s recommendations. Such evaluation should assess not only accuracy but also calibration, explanation quality, and failure mode characteristics. Critically, we find that these systems are not ready for autonomous decision-making. Therefore, applications should be designed with “human-in-the-loop” interactions and focus on usability for care providers.

Our hypotheses about why overthinking occurs and why P2 is robust to it remain speculative. Mechanistic interpretability studies examining model internals during reasoning—attention patterns, feature activations, and information flow—could provide deeper understanding of these phenomena and suggest principled interventions.

Importantly, PCCR can be combined with self-consistency where multiple PCCR responses can be sampled for the same question—voting over these multiple response could reduce overthinking and provide confidence measures for PCCR.

Our framework identifies equivalent configurations but does not optimize for hardware-specific efficiency. Combining our accuracy models with detailed latency and throughput modeling for specific deployment targets (e.g., consumer GPUs, edge devices, cloud instances) would enable more precise configuration recommendations.

Our evaluation focused on single-turn question answering. Clinical applications increasingly involve multi-turn dialogue and agentic workflows where models take actions based on reasoning. Whether our findings about prompting strategies generalize to these more complex settings remains an open question.

## 4 Conclusion

Deploying large language models for clinical reasoning requires balancing accuracy against the practical constraints of latency, cost, security, and accessibility. This study systematically evaluated the trade-offs between model scale, medical fine-tuning, test-time scaling, and context grounding using the Gemma and MedGemma model families across three medical question-answering datasets. We use our empirical results and observations to formulate a grounded framework for medical LLM system design. We identified an actionable bifurcation point in system design determined by whether high-quality context is available with clear implications for cost, accuracy, and latency. Our results point towards adaptive AI systems that modulate reasoning strategy based on context quality and availability to meet user objectives and minimize cost. Several limitations temper our conclusions and point towards necessary future work. Despite these limitations, our work provides a principled approach to navigating the trade-off space in clinical LLM system design and deployment. Our results demonstrate that current open-weight medical LLMs, even with optimal prompting configurations, do not achieve physician-level performance on novel clinical content. The best-performing configuration (MedGemma 27B with PCCR-1024) was below the 65% passing threshold and within the lowest quartile of practicing physicians. This gap persists despite the same configuration achieving 87.7% on established benchmarks like MedQA, suggesting that reported benchmark performance may substantially overestimate real-world capability on genuinely novel clinical reasoning tasks. Healthcare organizations should interpret published benchmark results cautiously and conduct institution-specific validation before deployment. As healthcare organizations increasingly consider adopting LLM-based tools across their clinical workflows, such rigorous approaches to system design will be a strong starting point and essential for realizing the promise of clinical AI.

## Data Availability

All data and code is available through a Github repo: https://github.com/MiningMyBusiness/accuracy-econ-medical-llms. This link is also provided directly in the manuscript text.

## A Appendix: Prompting strategies

### A.1 P0: Direct Answering

You are a medical expert. Answer the following multiple-choice question by selecting the correct option. Provide only the letter of your answer inside tags.

Question: A 45-year-old woman presents with fatigue, weight gain, and cold intolerance. Laboratory studies show elevated TSH and low free T4. Which of the following is the most likely diagnosis?

Graves’ disease
Hashimoto’s thyroiditis
Subacute thyroiditis
Toxic multinodular goiter

### A.2 P1: Chain-of-Thought

You are a medical expert. Answer the following multiple-choice question. First, reason through the problem step-by-step inside tags. Then, provide your final answer inside tags.

Question: A 45-year-old woman presents with fatigue, weight gain, and cold intolerance. Laboratory studies show elevated TSH and low free T4. Which of the following is the most likely diagnosis?

Graves’ disease
Hashimoto’s thyroiditis
Subacute thyroiditis
Toxic multinodular goiter

<think>

[Reason through the clinical presentation, laboratory findings, and each answer choice step-by-step]

</think>

### A.3 P2: Self-Consistency with Majority Voting

The P2 strategy uses the same prompt as P1, executed five times independently at temperature 0.7.

### A.4 PCCR: Prompt-Chaining for Continuous Reflection

The initial prompt is the same prompt as P1. Below we provide the continuation prompt. The continuation prompt is repeated until the cumulative token count within ¡think¿¡/think¿ tags meets or exceeds the specified budget (128, 256, 512, 768, or 1024 tokens). The final answer is extracted from the terminal response.

You are a medical expert continuing to reason through a multiple-choice question. Below is the original question and your reasoning so far. Continue reflecting on the problem: consider alternative interpretations, verify your logic, evaluate differential diagnoses you may have overlooked, and ensure your reasoning is complete. Add your continued reasoning inside tags, then provide your final answer inside tags.

Question: A 45-year-old woman presents with fatigue, weight gain, and cold intolerance. Laboratory studies show elevated TSH and low free T4. Which of the following is the most likely diagnosis?

Graves’ disease
Hashimoto’s thyroiditis
Subacute thyroiditis
Toxic multinodular goiter

Previous reasoning:

{PREVIOUS_REASONING}

Continue your analysis:

<think>

[Continue reasoning, verify conclusions, consider alternatives]

</think>

## B Appendix: LLM-as-judge prompt

You are an expert evaluator assessing whether a model’s answer to a multiple-choice medical question is correct. Your task is to determine if the model’s response matches the ground truth answer.

The model may have provided its answer in different formats:

– A single letter (e.g., “B”)
– A letter with explanation (e.g., “B) Hashimoto’s thyroiditis”)
– The full text of the answer choice
– An explanation followed by a final answer

You must determine whether the model’s final answer is factually equivalent to the ground truth answer. Focus on the model’s final/conclusive answer, not intermediate reasoning.

\## Evaluation Criteria

1. **Correct**: The model’s final answer matches the ground truth answer. This includes:

– Exact letter match (e.g., model says “B”, ground truth is “B”)
– Text match (e.g., model says “Hashimoto’s thyroiditis”, ground truth is “B) Hashimoto’s thyroiditis”)
– Semantically equivalent answers that clearly indicate the same choice
2. **Incorrect**: The model’s final answer does not match the ground truth, including:

– Different letter or answer choice selected
– Model failed to provide a clear final answer
– Model selected multiple answers when only one was requested
– Model explicitly stated it cannot answer or is uncertain without committing to a choice

\## Few-Shot Examples ### Example 1

Question: A 65-year-old man presents with crushing chest pain radiating to the left arm. ECG shows ST-elevation in leads V1-V4. Which is the most likely diagnosis?

Stable angina
Acute anterior STEMI
Pericarditis
Aortic dissection Ground Truth: B

Model Response: “The patient presents with classic symptoms of acute myocardial infarction - crushing chest pain with radiation and ST-elevation in the anterior leads (V1-V4). This pattern is consistent with an acute anterior STEMI. The answer is B) Acute anterior STEMI.”

Evaluation: <judgment>correct</judgment>

Reasoning: The model clearly selected option B, which matches the ground truth.

\### Example 2

Question: Which medication is first-line treatment for type 2 diabetes in patients without contraindications?

Insulin glargine
Metformin
Glipizide
Sitagliptin Ground Truth: B

Model Response: “For type 2 diabetes, we need to consider several factors. Metformin is generally recommended as first-line, but sulfonylureas like glipizide are also effective. Given the patient has no contraindications mentioned, I would consider either metformin or glipizide as reasonable options.”

Evaluation: <judgment>incorrect</judgment>

Reasoning: The model did not commit to a single final answer. While metformin was mentioned, the model equivocated between B and C without providing a definitive selection.

\### Example 3

Question: A 30-year-old woman presents with tremor, heat intolerance, and weight loss. Lab shows suppressed TSH and elevated free T4.

What is the most likely diagnosis?

A) Hashimoto’s thyroiditis
B) Graves’ disease
C) Toxic adenoma
D) Subacute thyroiditis Ground Truth: B

Model Response: “Graves’ disease”

Evaluation: <judgment>correct</judgment>

Reasoning: The model’s response “Graves’ disease” matches option B, the ground truth answer.

\### Example 4

Question: What is the most common cause of community-acquired pneumonia in adults?

A) Staphylococcus aureus
B) Klebsiella pneumoniae
C) Streptococcus pneumoniae
D) Pseudomonas aeruginosa Ground Truth: C

Model Response: “The most common cause is Staphylococcus aureus, which is responsible for the majority of community-acquired pneumonia cases. Answer: A”

Evaluation: <judgment>incorrect</judgment>

Reasoning: The model selected option A (Staphylococcus aureus), but the ground truth is C (Streptococcus pneumoniae).

---

\## Your Evaluation Task

Now evaluate the following: Question: {QUESTION}

Answer Choices:

{ANSWER_CHOICES}

Ground Truth: {GROUND_TRUTH}

Model Response: {MODEL_RESPONSE}

First, identify the model’s final answer. Then determine if it matches the ground truth. Provide brief reasoning followed by your judgment.

Reasoning: [Your reasoning here]

<judgment>[correct/incorrect]</judgment>

## C Appendix: Throughput Estimation Methodology

To estimate inference throughput for memory-bound decoding (batch size = 1), we utilized the Roofline model. For large language models, the generation phase is predominantly limited by memory bandwidth rather than compute capability. The theoretical maximum throughput *R* (tokens/second) is calculated as:

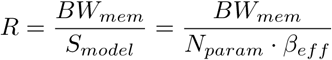

where:

- *BW_mem_* is the effective memory bandwidth of the hardware (GB/s).
- *N_param_* is the model parameter count (billions).
- *β_eff_* is the effective size per parameter in bytes, accounting for precision and quantization overhead.

### Precision Assumptions

We assumed the following effective memory footprints:

- **FP16 (Half Precision):** *β_eff_* = 2.0 bytes/parameter.
- **INT4 (Quantized):** *β_eff_* ≈ 0.65 bytes/parameter (0.5 bytes for 4-bit weights + 0.15 bytes for group-wise quantization scales and zeros).

### Hardware Specifications

Bandwidth values (*BW_mem_*) were set as follows:

- NVIDIA A100 (80GB SXM4): 1,935 GB/s.
- NVIDIA RTX 4090: 1,008 GB/s.
- Consumer CPU (DDR5 Dual-Channel): ∼60 GB/s (estimated effective bandwidth for DDR5-6000).

### Latency Bounds

For smaller models (e.g., 1B), the theoretical memory-bound throughput often exceeds the kernel launch latency limit. In these regimes, realized throughput is typically capped by compute overhead (kernel latency), often plateauing around 200–300 tokens/s on current GPU architectures regardless of available bandwidth.

## D Appendix: Query Volume Estimation

To accurately amortize the fixed costs of fine-tuning, we estimate the annual query volume *Q_life_* for a representative large academic health system. We assume the model is integrated into the electronic health record (EHR) workflow to assist with clinical documentation and decision support for every patient encounter.

### Operational Baseline

We model a large integrated health network based on typical metrics for top-tier US hospital systems (e.g., systems with *>*2,000 beds):

- **Clinical Staff (***N_staff_* ): 4,500 active providers (attending physicians, residents, and advanced practice providers).
- **Patient Encounters (***R_enc_*): Average of 18 encounters per provider per workday (outpatient visits, inpatient rounds, or procedure notes).
- **Operational Days (***D_ops_*): 230 clinical workdays per year per provider.

### Query Generation Rate

We define the Query Rate (*λ*) as the number of model calls triggered per encounter. For a system providing automated note drafting or background clinical decision support (CDS), *λ* ≥ 1.

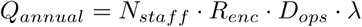

Table 21 calculates the projected volumes.

**Table 21.** Estimated annual query volume for a large hospital system.

| Parameter | Value | Rationale |
| --- | --- | --- |
| Clinical Providers ($N_{staff}$) | 4,500 | Multi-hospital system (2,000+ beds) |
| Encounters/Day ($R_{enc}$) | 18 | Avg. combined inpatient/outpatient load |
| Workdays/Year ($D_{ops}$) | 230 | Accounting for vacation/admin time |
| **Total Encounters** | **18,630,000** | Baseline opportunity volume |
| Query Rate ($\lambda$) | 1.0 | 1 query per encounter (e.g., Note Draft) |
| **Annual Query Volume** | **~18.6 Million** | $Q_{annual}$ |

We approximate *20 million queries per year* as a reference query volume based on reasonable approximations. However, these estimates consider use at-scale with every encounter. Actual values during pilot phases or before broad adoption could be lower. Alternatively, multiple such systems serving different use-cases could result in multiple uses per encounter, leading to linear increases in query volume. Agentic orchestrations where services can query each other, could lead to far greater consumption.

## E Appendix: RAG Cost Estimation Methodology

To determine a realistic retrieval cost per query (*C_RAG_*), we utilize a Total Cost of Ownership (TCO) approach that distinguishes between marginal variable costs (OpEx) and amortized fixed infrastructure costs (CapEx + OpEx).

The total cost per query is modeled as:

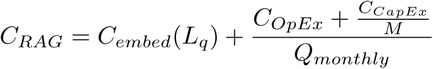

where:

- *L_q_* is the average query length in tokens.
- *Q_monthly_* is the total number of queries processed per month.
- *M* is the amortization period in months (typically 36 for hardware).

1. Variable Embedding Cost (*C_embed_*). This represents the API cost to embed the user’s query vector. where *P_emb_* is the price per million tokens (e.g., ∼ $0.02 for efficient embedding models). For a typical query (*L_q_* ≈ 100), this cost is negligible (≈ $0.000002).

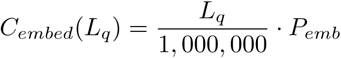
2. Fixed Operational Expenses (*C_OpEx_*). This includes monthly recurring costs for the vector database and retrieval infrastructure: *C_OpEx_* = *P_cloud db_* + *P_hosting_* Example: A production-grade managed vector database (e.g., Pinecone, Weaviate) typically costs $50–$150/month for moderate distinct vector counts (*<* 1*M*).

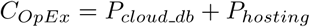
3. Capital Expenses (*C_CapEx_*). For on-premise or private cloud deployments utilizing dedicated hardware (e.g., GPU servers for local re-ranking or embedding): *C_CapEx_* = Hardware Cost + Initial Setup Cost

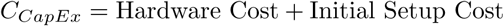

### Sensitivity to Scale

RAG costs are dominated by vector database and GPU server cost for hosting embedding models. As shown below, low-volume applications face significantly higher unit costs due to unamortized fixed infrastructure:

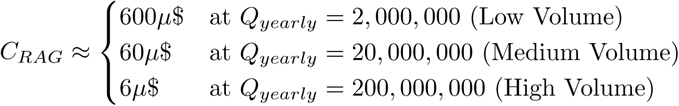

Values shown in micro-dollars (*µ*$ = 10^−6^ USD). At the expected query volume of 20 million queries per year, we should expect an amortized cost per query of 60*µ*$ or $0.00006. We round this up to $0.0001 as the reference cost for our calculations.

## F Appendix: Fine-tuning Cost Estimation

The total capital expenditure for fine-tuning (*C_F T_*) is the sum of compute costs and data acquisition costs:

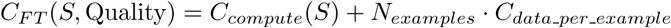

### Compute Costs

Based on parameter-efficient fine-tuning (LoRA/QLoRA) benchmarks, compute costs are relatively minor compared to high-quality data costs. For a 27B model, fine-tuning requires approximately 4–12 hours on a single H100 or 8–24 hours on dual A100s, costing roughly $40–$192.

### Data Costs

Data costs vary by orders of magnitude depending on the source:

- Synthetic (LLM-generated): $0.01–0.05 per example.
- Curated public/private: $0.10–0.50 per example (cleaning/formatting/annotation).
- Expert-annotated (Clinical): $5–50 per example (physician or subject-matter-expert review).

Table 22 summarizes the total estimated fine-tuning costs.

**Table 22.**
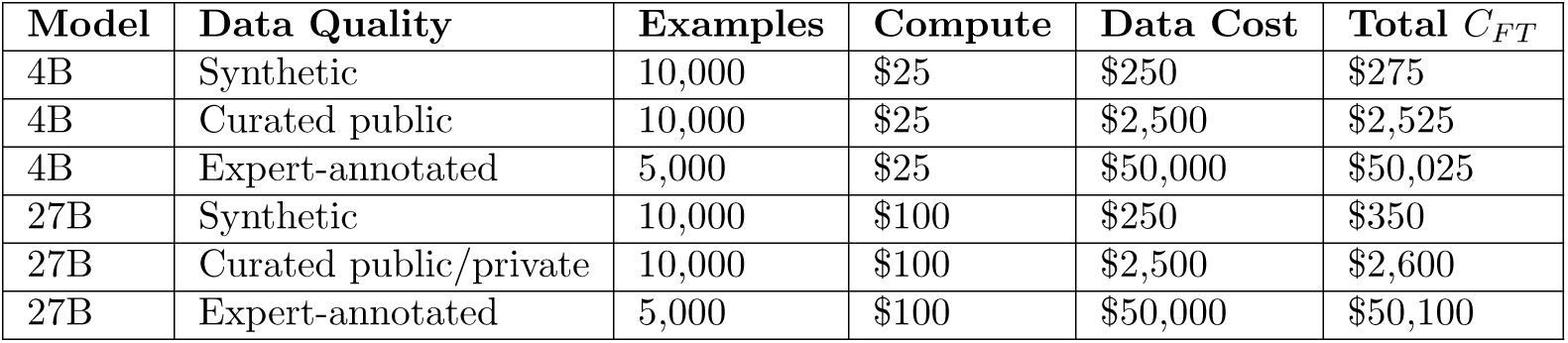
Estimated total fine-tuning costs (*C_F T_*) by model size and data quality.

### Amortization

The impact of *C_F T_* on per-query cost depends heavily on query volume over the lifetime of the model (*Q_life_*). Table 23 presents the amortized cost added to each query.

**Table 23.** Amortized fine-tuning cost per query by volume and data quality for 27B model. Values shown in micro-dollars (*µ*$ = 10^−6^ USD). e.g., 35, 000*µ*$ = $0.035.

| Query Volume | Synthetic (\$350) | Curated (\$2,600) | Expert (\$50,100) |
| --- | --- | --- | --- |
| 20,000 | $17,500\mu\$$ | $130,000\mu\$$ | $2,505,000\mu\$$ (\$5.01) |
| 200,000 | $1,750\mu\$$ | $13,000\mu\$$ | $250,500\mu\$$ (\$0.50) |
| 2,000,000 | $175\mu\$$ | $1,300\mu\$$ | $25,050\mu\$$ (\$0.05) |
| 20,000,000 | $17.5\mu\$$ | $130\mu\$$ | $2,505\mu\$$ (\$0.005) |
| 200,000,000 | $1.75\mu\$$ | $13\mu\$$ | $250.5\mu\$$ (\$0.0005) |

At low volumes (*<*100k queries), expert-annotated fine-tuning adds significant cost ($0.50–$5.00 per query). At the reference query volume of 20 million queries per year, the amortized cost per query has a large range from 17.5*µ*$ − 2, 505*µ*$ based on data quality. Assuming that synthetic and curated dataset based fine-tuning can be performed by vendors, hospitals may only need to fine-tune on high-quality, expert-reviewed data. However, in this study, we use off-the-shelf fine-tuned models trained on curated data and, likely, synthetic data. Therefore, we choose the middle of the range as a reasonable per query amortization cost ∼ 1250*µ*$ or $0.00125. We round this to $0.0013 for convenience.

## G Appendix: More example solutions to the optimization problem

### G.0.1 Example 3: Latency-critical application

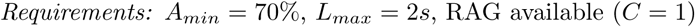

We enumerate candidate configurations from Table 16 with *C* = 1 and achieving ≥ 70% to populate Table 24. The optimal solution configuration is (4, 0, P2, 1) or Gemma-3 4B + P2 + RAG at 145*µ*$ per query and a latency of 1.8 seconds. Note that this latency can be reduced further and the accuracy improved by selecting the MedGemma 4B fine-tuned model, if the higher cost is still palatable.

**Table 24.** Candidate configurations for Example 3: Latency-critical application that meet the accuracy requirement. Values shown in micro-dollars (*µ*$ = 10^−6^ USD).

| Config | Accuracy | Cost | Latency | Feasible? |
| --- | --- | --- | --- | --- |
| (27, 1, P0, 1) | 74.7% | 1412$\mu\$$ | 3.7s | No (latency) |
| (27, 1, P1, 1) | 77.4% | 1430$\mu\$$ | 8.9s | No (latency) |
| (27, 0, P2, 1) | 70.1% | 250$\mu\$$ | 8.9s | No (latency) |
| (4, 1, P0, 1) | 73.1% | 1403$\mu\$$ | 0.9s | **Yes** |
| (4, 0, P2, 1) | 70.1% | 145$\mu\$$ | 1.8s | **Yes** |
| (4, 0, PCCR-128, 1) | 76.0% | 114.76$\mu\$$ | 2.76s | No (latency) |

### G.0.2 Example 4: Budget-constrained application

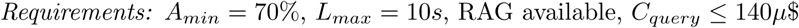

We enumerate candidate configurations from Table 16 with *C* = 1 and achieving ≥ 70% to populate Table 25. The optimal solution configuration is (4, 0, PCCR-128, 1) or

**Table 25.** Candidate configurations for Example 4: Latency-critical application that meet the accuracy requirement. Values shown in micro-dollars (*µ*$ = 10^−6^ USD).

| Config | Accuracy | Cost | Latency | Feasible? |
| --- | --- | --- | --- | --- |
| (27, 1, P0, 1) | 74.7% | 1412$\mu\$$ | 3.7s | No (cost) |
| (27, 1, P1, 1) | 77.4% | 1430$\mu\$$ | 8.9s | No (cost) |
| (27, 0, P2, 1) | 70.1% | 250$\mu\$$ | 8.9s | No (cost) |
| (4, 1, P0, 1) | 73.1% | 1403$\mu\$$ | 0.9s | No (cost) |
| (4, 0, P2, 1) | 70.1% | 145$\mu\$$ | 1.8s | No (cost) |
| (4, 0, PCCR-128, 1) | 76.0% | 114.76$\mu\$$ | 2.76s | **Yes** |

Gemma-3 4B + PCCR-128 + RAG at 114*µ*$ per query and a latency of 2.76 seconds. Low-cost requirements will favor generic models, as can be inferred from Figure 4.

## References

1. Nori H, Lee YT, Zhang S, Carignan D, Edgar R, Fusi N, et al. Can generalist foundation models outcompete special-purpose tuning? case study in medicine. arXiv preprint arXiv:231116452. 2023;.

2. Saab K, Tu T, Weng WH, Tanno R, Stutz D, Wulczyn E, et al. Capabilities of gemini models in medicine. arXiv preprint arXiv:240418416. 2024;.

3. Dennstädt F, Hastings J, Putora PM, Schmerder M, Cihoric N. Implementing large language models in healthcare while balancing control, collaboration, costs and security. NPJ digital medicine. 2025;8(1):143.

4. Kaplan J, McCandlish S, Henighan T, Brown TB, Chess B, Child R, et al. Scaling laws for neural language models. arXiv preprint arXiv:200108361. 2020;.

5. Hoffmann J, Borgeaud S, Mensch A, Buchatskaya E, Cai T, Rutherford E, et al. Training compute-optimal large language models (2022). arXiv preprint arXiv:220315556. 2022;.

6. Singhal K, Azizi S, Tu T, Mahdavi SS, Wei J, Chung HW, et al. Large language models encode clinical knowledge. Nature. 2023;620(7972):172–180. doi:10.1038/s41586-023-06291-2.

7. Ke YH, Jin L, Elangovan K, Abdullah HR, Liu N, Sia ATH, et al. Retrieval augmented generation for 10 large language models and its generalizability in assessing medical fitness. npj Digital Medicine. 2025;8(1):187.

8. Ng KKY, Matsuba I, Zhang PC. RAG in health care: a novel framework for improving communication and decision-making by addressing LLM limitations. NEJM AI. 2025;2(1):AIra2400380.

9. Shi Y, Xu S, Yang T, Liu Z, Liu T, Li X, et al. Mkrag: Medical knowledge retrieval augmented generation for medical question answering. In: AMIA Annual Symposium Proceedings. vol. 2024; 2025. p. 1011.

10. Zhang H, et al. Discuss-RAG: Enhancing Retrieval-Augmented Generation via Agent-Led Multi-Turn Reasoning for Medical QA. arXiv preprint arXiv:250421252. 2024;.

11. Xu K, Zhang K, Li J, Huang W, Wang Y. CRP-RAG: A Retrieval-Augmented Generation Framework for Supporting Complex Logical Reasoning and Knowledge Planning. Electronics. 2024;14(1):47.

12. Liu S, McCoy AB, Wright A. Improving large language model applications in biomedicine with retrieval-augmented generation: a systematic review, meta-analysis, and clinical development guidelines. Journal of the American Medical Informatics Association. 2025; p. ocaf008.

13. Hu EJ, Shen Y, Wallis P, Allen-Zhu Z, Li Y, Wang S, et al. Lora: Low-rank adaptation of large language models. ICLR. 2022;1(2):3.

14. Chen Z, Cano AH, Romanou A, Bonnet A, Matoba K, Salvi F, et al. Meditron-70b: Scaling medical pretraining for large language models. arXiv preprint arXiv:231116079. 2023;.

15. Sellergren A, Kazemzadeh S, Jaroensri T, Kiraly A, Traverse M, Kohlberger T, et al. Medgemma technical report. arXiv preprint arXiv:250705201. 2025;.

16. Thirunavukarasu AJ, Ting DSJ, Elangovan K, Gutierrez L, Tan TF, Ting DSW. Large language models in medicine. Nature medicine. 2023;29(8):1930–1940.

17. Huang X, Wu J, Liu H, Tang X, Zhou Y. m1: Unleash the potential of test-time scaling for medical reasoning with large language models. arXiv preprint arXiv:250400869. 2025;.

18. Muennighoff N, Yang Z, Shi W, Li XL, Fei-Fei L, Hajishirzi H, et al. s1: Simple test-time scaling. In: Proceedings of the 2025 Conference on Empirical Methods in Natural Language Processing; 2025. p. 20286–20332.

19. Snell C, Lee J, Xu K, Kumar A. Scaling llm test-time compute optimally can be more effective than scaling model parameters. arXiv preprint arXiv:240803314. 2024;.

20. Team G, Kamath A, Ferret J, Pathak S, Vieillard N, Merhej R, et al. Gemma 3 technical report. arXiv preprint arXiv:250319786. 2025;.

21. Xiong G, Jin Q, Lu Z, Zhang A. Benchmarking retrieval-augmented generation for medicine. In: Findings of the Association for Computational Linguistics ACL 2024; 2024. p. 6233–6251.

22. Jin Q, Dhingra B, Liu Z, Cohen W, Lu X. PubMedQA: A Dataset for Biomedical Research Question Answering. In: Proceedings of the 2019 Conference on Empirical Methods in Natural Language Processing and the 9th International Joint Conference on Natural Language Processing (EMNLP-IJCNLP); 2019. p. 2201–2211.

23. Katz U, Cohen E, Shachar E, Somer J, Fink A, Morse E, et al. GPT versus resident physicians—a benchmark based on official board scores. Nejm Ai. 2024;1(5):AIdbp2300192.

24. Wei J, Wang X, Schuurmans D, Bosma M, Xia F, Chi E, et al. Chain-of-thought prompting elicits reasoning in large language models. Advances in neural information processing systems. 2022;35:24824–24837.

25. Kojima T, Gu SS, Reid M, Matsuo Y, Iwasawa Y. Large language models are zero-shot reasoners. Advances in neural information processing systems. 2022;35:22199–22213.

26. Wang X, Wei J, Schuurmans D, Le Q, Chi E, Narang S, et al. Self-consistency improves chain of thought reasoning in language models. arXiv preprint arXiv:220311171. 2022;.

27. Chen X, Aksitov R, Alon U, Ren J, Xiao K, Yin P, et al. Universal self-consistency for large language model generation. arXiv preprint arXiv:231117311. 2023;.

28. Savage T, Wang J, Gallo R, Boukil A, Patel V, Safavi-Naini SAA, et al. Large language model uncertainty proxies: discrimination and calibration for medical diagnosis and treatment. Journal of the American Medical Informatics Association. 2025;32(1):139–149.

29. Wolf T, Debut L, Sanh V, Chaumond J, Delangue C, Moi A, et al. Transformers: State-of-the-art natural language processing. In: Proceedings of the 2020 conference on empirical methods in natural language processing: system demonstrations; 2020. p. 38–45.

30. Jain AM, Jindal M. Systematic survey of various prompt optimization methods and their classifications. In: 2025 11th International Conference on Computing and Artificial Intelligence (ICCAI). IEEE; 2025. p. 524–536.

31. Yao S, Yu D, Zhao J, Shafran I, Griffiths TL, Cao Y, et al. Tree of thoughts: Deliberate problem solving with large language models, 2023. URL https://arxiv org/abs/230510601. 2023;3:1.

32. Madaan A, Tandon N, Gupta P, Hallinan S, Gao L, Wiegreffe S, et al. Self-refine: Iterative refinement with self-feedback. Advances in Neural Information Processing Systems. 2023;36:46534–46594.

33. Shinn N, Cassano F, Gopinath A, Narasimhan K, Yao S. Reflexion: Language agents with verbal reinforcement learning. Advances in Neural Information Processing Systems. 2023;36:8634–8652.

34. Du Y, Li S, Torralba A, Tenenbaum JB, Mordatch I. Improving factuality and reasoning in language models through multiagent debate. In: Forty-first International Conference on Machine Learning; 2023.

35. Liang T, He Z, Jiao W, Wang X, Wang Y, Wang R, et al. Encouraging divergent thinking in large language models through multi-agent debate. In: Proceedings of the 2024 conference on empirical methods in natural language processing; 2024. p. 17889–17904.

36. Xiong G, Jin Q, Wang X, Zhang M, Lu Z, Zhang A. Improving retrieval-augmented generation in medicine with iterative follow-up questions. In: Biocomputing 2025: Proceedings of the Pacific Symposium. World Scientific; 2024. p. 199–214.

37. Chen J, Cai Z, Ji K, Wang X, Liu W, Wang R, et al. Towards medical complex reasoning with LLMs through medical verifiable problems. In: Findings of the Association for Computational Linguistics: ACL 2025; 2025. p. 14552–14573.

